# Distribution and associated factors of hepatic iron – a population-based imaging study

**DOI:** 10.1101/2021.10.11.21264730

**Authors:** Lisa Maier, Ricarda von Krüchten, Roberto Lorbeer, Jule Filler, Johanna Nattenmüller, Barbara Thorand, Wolfgang Koenig, Wolfgang Rathmann, Fabian Bamberg, Christopher L. Schlett, Annette Peters, Susanne Rospleszcz

**Author notes:** **Correspondence and Reprint Requests:** Susanne Rospleszcz, Institute of Epidemiology, Helmholtz Zentrum München, German Research Center for Environmental Health, Ingolstädter Landstraße 1, 85764 Neuherberg, Germany.

## Abstract

**Context:** Hepatic iron overload can cause severe organ damage. Therefore, an early diagnosis is crucial, and identification of modifiable risk factors could help to prevent manifestations of iron-driven complications.

**Objective:** To investigate the sex-specific distribution of hepatic iron content (HIC) in a population-based sample, and to identify relevant associated factors from a panel of markers.

**Methods:** We analysed N=353 participants from a cross-sectional, population-based cohort in Southern Germany (KORA FF4) who underwent whole-body magnetic resonance imaging. HIC was assessed by single-voxel spectroscopy with a high-speed T2-corrected multi-echo technique. A large panel of markers, including anthropometric, genetic and laboratory values as well as behavioural risk factors were assessed. Relevant factors associated with HIC were identified by variable selection based on LASSO regression with bootstrap resampling.

**Results:** HIC in the study sample (mean age at examination was 56.0 years, 58.4% were men) was significantly lower in women (mean±SD: 39.2±4.1 s^-1^) than in men (41.8±4.7 s^-1^, p<0.001). Relevant factors associated with HIC were HbA1c and prediabetes for men, and visceral adipose tissue and age for women. Hepatic fat, alcohol consumption, and a genetic risk score for iron levels were associated with HIC in both sexes.

**Conclusion:** There are sex-specific associations of HIC with markers of body composition, glucose metabolism and alcohol consumption.

## Introduction

Iron is an essential element in human organisms. It is of great importance for the transport and storage of oxygen, but also regulates cell survival and DNA synthesis.

Consequently, deviations from normal ranges of stored body iron are associated with the development of certain pathologies. Excess in body iron storage leads to potential cell damage due to the generation of reactive oxygen species (ROS). These highly reactive oxygens induce lipid peroxidation and DNA damage resulting, among others, in liver injuries (1).

In particular, the liver plays an important role in maintaining iron homeostasis. Hepcidin, a protein regulated by the HAMP gene and expressed within the liver, is the main regulator of iron homeostasis. Its expression is stimulated in the presence of iron overload to inhibit the resorption of iron. Moreover, the liver is the main storage site of iron and is susceptible to iron overload due to iron accumulation in hepatocytes (2). Hepatic iron content (HIC) serves as a surrogate for whole-body iron storage (3). Excessive hepatic iron storage can progress to severe liver diseases, such as fibrosis, cirrhosis, or hepatocellular carcinoma (4).

Mechanisms responsible for the disruption of iron homeostasis and pathways associated with comorbidities are still insufficiently explored. However, studies have linked increased HIC with type 2 diabetes mellitus (T2DM) and insulin resistance (5), hypertension (6), and non-alcoholic fatty liver disease (NAFLD) (2) suggesting a cross-talk between metabolic syndrome (MetS) and iron metabolism.

HIC increases gradually to pathological levels (7). Therefore, an early diagnosis of elevated HIC and identification of relevant, potentially modifiable risk factors would be beneficial to prevent manifestations of iron-driven organ damage and further complications.

However, clinical assessment of HIC is challenging. Early presentations of hepatic iron overload range from asymptomatic to mild cases or patients presenting with predominantly non-specific symptoms (8). Population-based studies are scarce since liver biopsy, the gold standard for HIC assessment, is an invasive procedure and not feasible at a population level. Hence, the majority of studies on HIC are based on small patient cohorts (9, 10). Alternatively, serum ferritin is regularly assessed as an indirect marker for body iron stores. Several population-based studies have already been conducted analysing associations of serum ferritin with metabolic disorders (11, 12). However, the interpretability of this biomarker is limited, as serum ferritin is also influenced by inflammation and coexisting liver diseases, and therefore might be artificially elevated (13).

Magnetic resonance imaging (MRI) has evolved as a powerful non-invasive diagnostic tool to accurately assess HIC. Nonetheless, only few studies have so far investigated the distribution of HIC in population-based samples and reported early evidence on a limited number of associated factors (14, 15).

Therefore, we aim to determine the sex-specific distribution of MRI-derived HIC in a population-based study, and to identify relevant associated factors form a broad panel of markers.

## Materials and Methods

### Study Design and Participants

The study sample consists of participants from the cross-sectional KORA MRI study (KORA: “Cooperative Health Research in the Region of Augsburg”), nested within the KORA FF4 study (N=2279, enrolled between 2013-2014). KORA FF4 is the second follow-up of the population-based KORA S4 cohort (N=4261, enrolled between 1999-2001). Overall, study design, recruitment and data collection of the KORA studies have been described in detail elsewhere (16). The KORA MRI sub-study includes 400 participants who underwent whole-body MRI, with a focus on assessing subclinical cardiometabolic diseases at different stages of impaired glucose metabolism (17). Briefly, participants with a history of cardiovascular disease, older than 73 years, or with any contraindications to whole-body MRI were excluded. For the current analysis, a total of 47 participants had to be excluded due to missing hepatic iron measurements or covariables, yielding a final main sample size of 353 participants. The detailed participant flow is shown in Figure 1.

**Figure 1:**
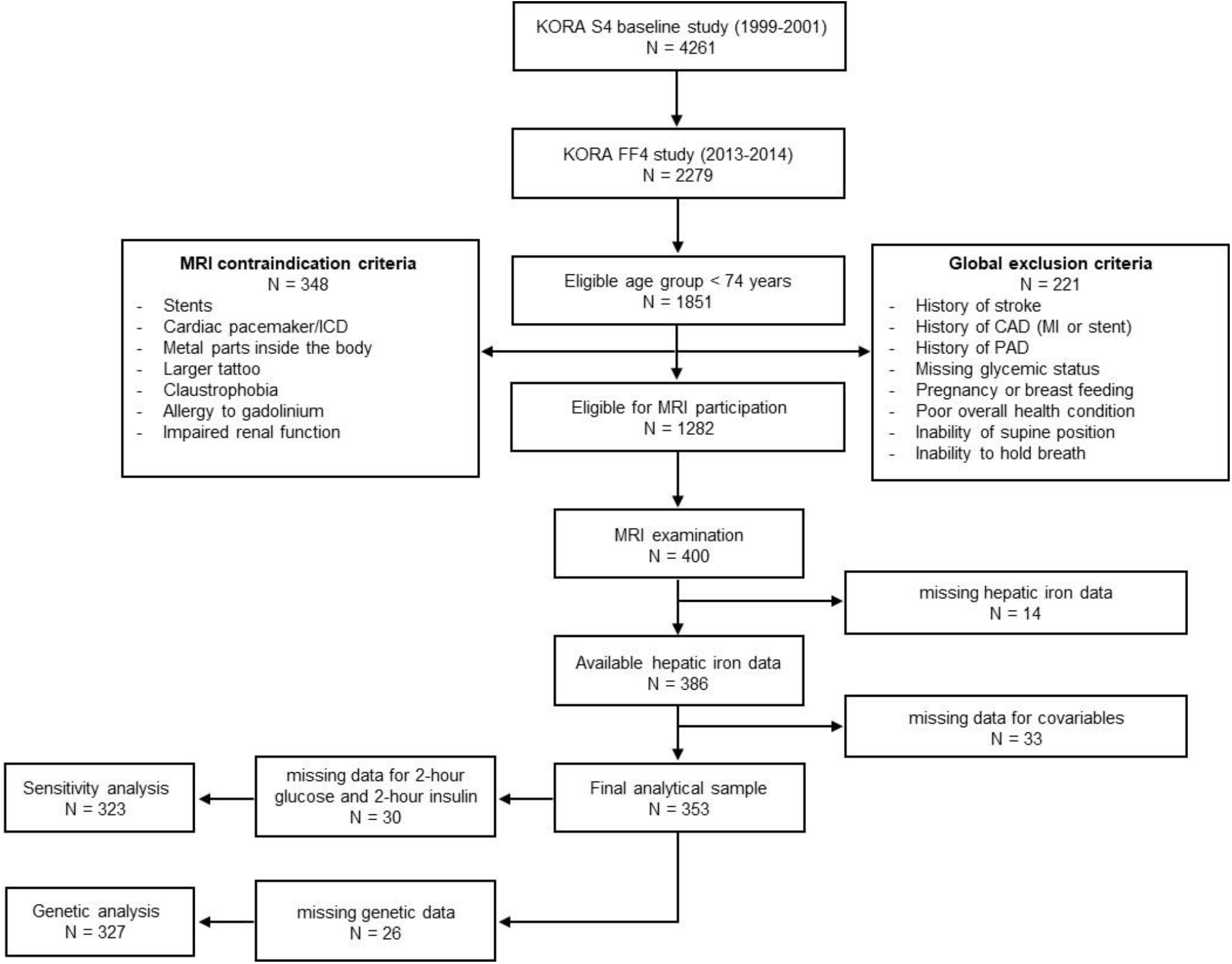
Participant flow chart. MRI, magnetic resonance imaging; CAD, coronary artery disease; MI, myocardial infarction; PAD, peripheral artery disease; ICD, implantable cardioverter defibrillator.

The study was approved by the ethics committee of the Ludwig-Maximilians-University Munich and the Bavarian Chamber of Physicians. It was performed according to the Declaration of Helsinki and all participants gave written informed consent.

### Outcome and Exposure Assessment

#### MRI examination: Hepatic iron and fat content

MRI examinations were performed on a 3 Tesla MRI scanner (Magnetom Skyra; Siemens AG, Siemens Healthineers, Erlangen, Germany). The protocol comprised dedicated sequences for the respective body regions, as detailed elsewhere (17). HIC was measured in the right and left hepatic lobe (segments VII and II, respectively) using a single-voxel spectroscopy with a high-speed T2-corrected multi-echo (HISTO) technique, allowing for the simultaneous assessment of hepatic iron and hepatic fat (18). Hepatic fat was obtained as hepatic fat fraction (HFF) in percent and averaged over the left and right liver lobe. HIC was quantified as relaxation rate 1/T2^*^ in s^-1^ in the left and right liver lobe. The arithmetic mean of left and right lobe constitutes the main outcome of the present analysis.

#### Covariates

A set of health-related covariables was collected from all KORA FF4 participants at the study center in a standardized fashion. Briefly, the assessment comprised laboratory values, anthropometric measurements, information about medication intake, sociodemographic characteristics and health behaviour (e.g. smoking, physical activity). Data were collected and maintained by trained staff according to standardized protocols. A venous blood sample in fasted condition was drawn from each participant to determine laboratory values. The laboratory analysis included a standard complete blood count, blood lipids, glucose metabolism markers, renal function parameters, an electrolyte panel, and liver enzymes. Further information is detailed in Supplementary Table 1. Since 2-hour insulin and 2-hour glucose data were only available for participants without established T2DM, sensitivity analyses including these variables were performed on a smaller sample without participants with diagnosed T2DM.

Furthermore, visceral and subcutaneous adipose tissue (VAT and SAT) were measured by MRI using a three-dimensional in/opposed-phase VIBE-Dixon sequence from the femoral head to the diaphragm and cardiac apex, respectively. VAT and SAT were post-processed using an automated algorithm-driven procedure for segmentation (19) and are given in liter (l). Genotyping was done with the Affymetrix Axiom Chip (20) and subsequent imputation was based on the Haplotype Reference Consortium (HRC) imputation panel r1.1, resulting in post-imputation probabilities (dosages) per allele. A genetic risk score was calculated to estimate the combined effect of selected single-nucleotide polymorphisms (SNPs) on HIC. Relevant SNPs associated with markers of iron metabolism were identified by querying the GWAS Catalogue (https://www.ebi.ac.uk/gwas/).

SNPs were then weighted by coefficients from sex-stratified univariate linear regressions against HIC (Supplementary Table 2), multiplied by the respective allele dosage, and summed up.

### Statistical Analysis

Descriptive statistics for continuous variables are presented as either arithmetic mean and standard deviation (SD) or median and interquartile range (IQR), where appropriate, and as counts and percentages for categorical variables. Differences between male and female participants were tested using t-test, Mann-Whitney-U test or χ^2^ -test, respectively. Correlations between HIC and continuous exposure variables were determined by Spearman’s rho correlation coefficient and corresponding p-values. Additionally, participants were classified according to presence of hepatic steatosis defined by a cutoff of HFF ≥ 5.6 % (21). All analyses were stratified by sex.

To identify relevant factors associated with HIC, least absolute shrinkage and selection operator (LASSO) linear regression was performed. LASSO is particularly suitable for this exploratory study, as it constitutes a variable selection method able to extract the most strongly associated factors from a large set of potentially correlated variables (22). LASSO achieves variable selection by applying a regularization process, where regression coefficients of less associated variables are shrunk towards zero by adding a penalty term λ. To quantify the relative importance of the selected variables and assess model stability, 1000 bootstrap samples were generated, and the LASSO regression model was fitted on each bootstrap sample. The penalty term λ was optimized for each bootstrap sample via 10-fold cross-validation. The percentage of variable inclusion among the 1000 bootstrap samples was calculated to quantify the relative importance of each variable, and variables with inclusion frequencies > 20% were considered relevant (23). Due to the regularization procedure, LASSO coefficients are biased towards zero. Therefore, calculation of confidence intervals and p-values is not straightforward.

To assess the strength of associations between covariables and HIC, unpenalized linear regression analyses adjusted for age and HFF were applied for every variable selected in LASSO regression. Results from unpenalized regression analyses are reported as unstandardized beta coefficients with corresponding confidence intervals, p-values and adjusted R^2^. Variables with a highly skewed distribution were log-transformed before regression analyses.

To further assess model stability, both penalized LASSO regressions and unpenalized regressions were run excluding HFF as a covariate.

In this exploratory analysis, p-values were not corrected for multiple testing and values less than 0.05 were considered to indicate statistical significance. All analyses were performed using R version 3.6.1.

## Results

### Study sample

Characteristics of the study sample are provided in Table 1. Age at the time of examination was 56.0 ± 9.1 (mean ± SD) years, 58.4% were male. Among the 353 participants, 12.2% had diagnosed diabetes, 23.5% had prediabetes and 64.3% were normoglycemic. Men had significantly higher values of HFF than women (median (IQR): 7.02% (10.08) and 3.53% (4.28), p<0.001, respectively).

**Table 1:**
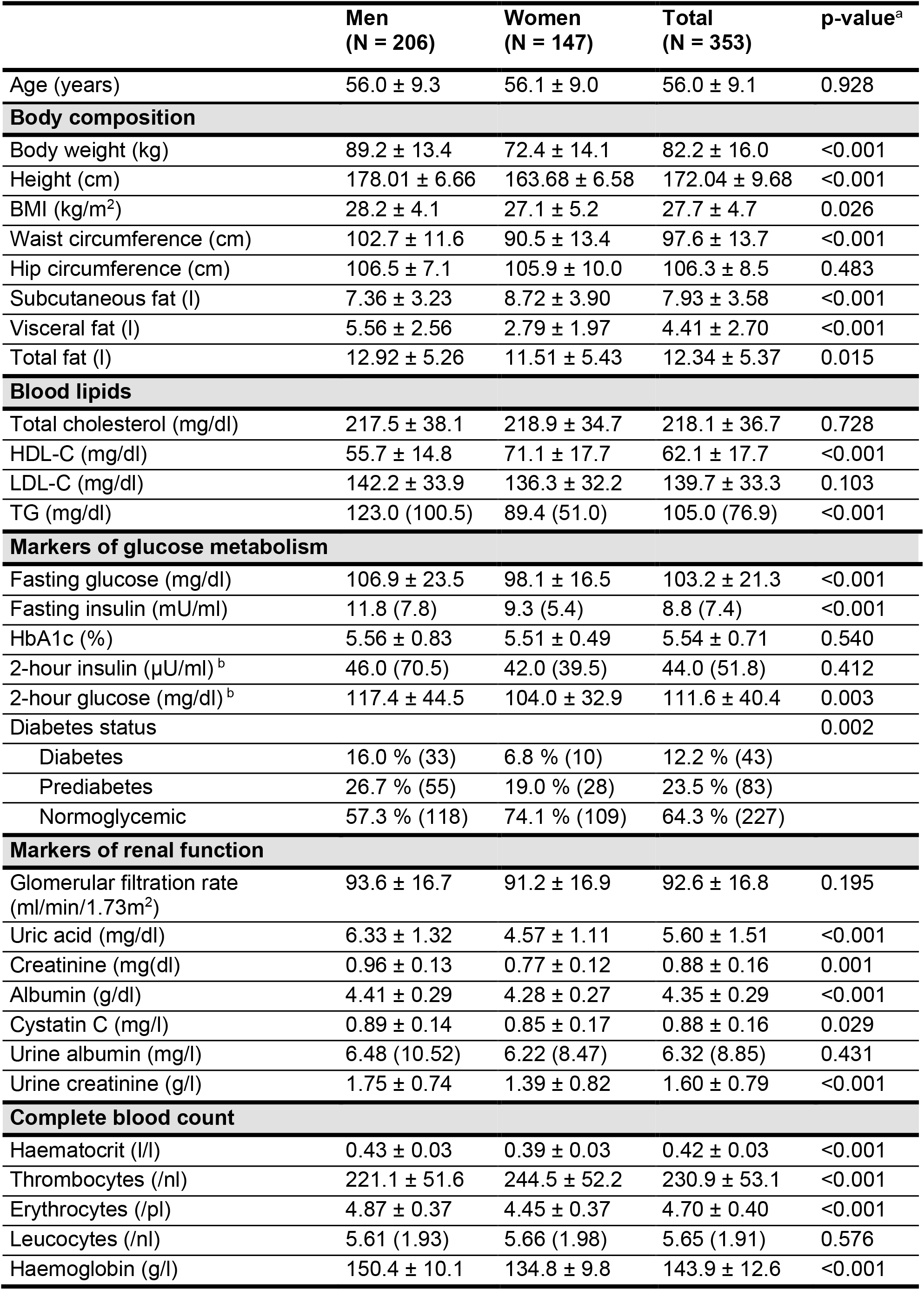
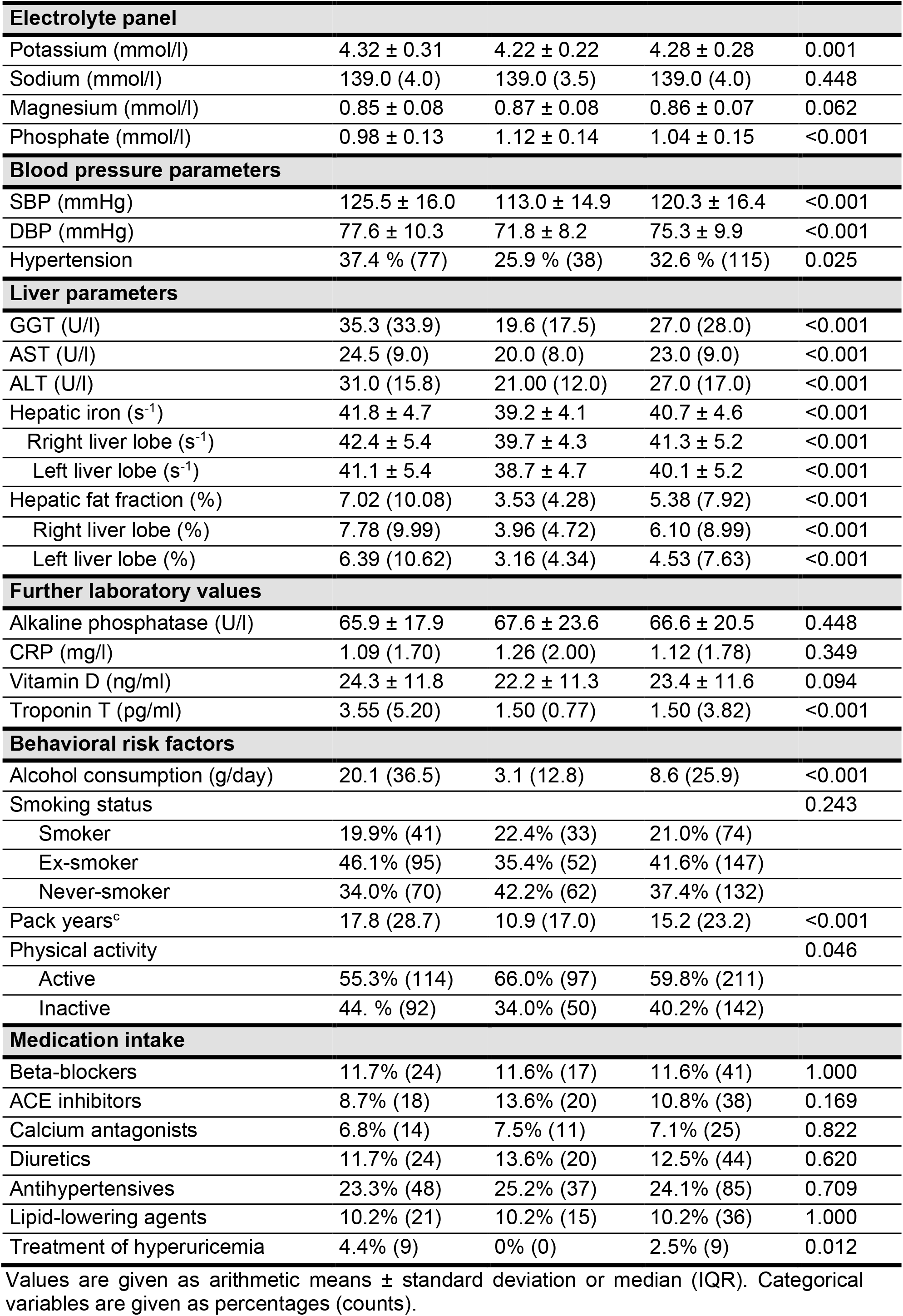
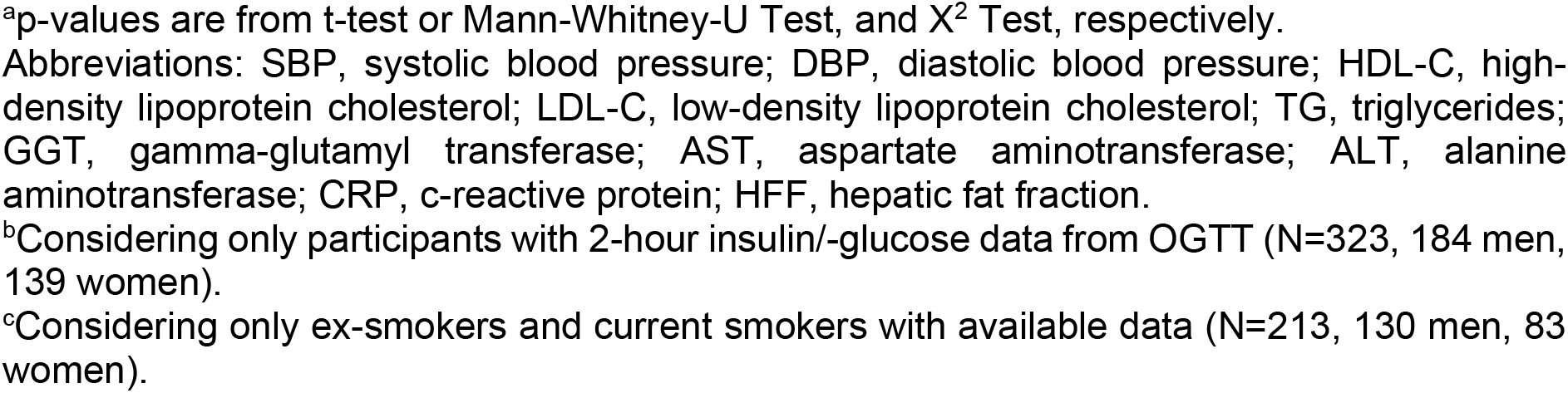
Descriptive characteristics of the study participants by sex.

|  | **Men**<br>**(N = 206)** | **Women**<br>**(N = 147)** | **Total**<br>**(N = 353)** | **p-value<sup>a</sup>** |
| --- | --- | --- | --- | --- |
| Age (years) | 56.0 ± 9.3 | 56.1 ± 9.0 | 56.0 ± 9.1 | 0.928 |
| **Body composition** |  |  |  |  |
| Body weight (kg) | 89.2 ± 13.4 | 72.4 ± 14.1 | 82.2 ± 16.0 | <0.001 |
| Height (cm) | 178.01 ± 6.66 | 163.68 ± 6.58 | 172.04 ± 9.68 | <0.001 |
| BMI (kg/m<sup>2</sup>) | 28.2 ± 4.1 | 27.1 ± 5.2 | 27.7 ± 4.7 | 0.026 |
| Waist circumference (cm) | 102.7 ± 11.6 | 90.5 ± 13.4 | 97.6 ± 13.7 | <0.001 |
| Hip circumference (cm) | 106.5 ± 7.1 | 105.9 ± 10.0 | 106.3 ± 8.5 | 0.483 |
| Subcutaneous fat (l) | 7.36 ± 3.23 | 8.72 ± 3.90 | 7.93 ± 3.58 | <0.001 |
| Visceral fat (l) | 5.56 ± 2.56 | 2.79 ± 1.97 | 4.41 ± 2.70 | <0.001 |
| Total fat (l) | 12.92 ± 5.26 | 11.51 ± 5.43 | 12.34 ± 5.37 | 0.015 |
| **Blood lipids** |  |  |  |  |
| Total cholesterol (mg/dl) | 217.5 ± 38.1 | 218.9 ± 34.7 | 218.1 ± 36.7 | 0.728 |
| HDL-C (mg/dl) | 55.7 ± 14.8 | 71.1 ± 17.7 | 62.1 ± 17.7 | <0.001 |
| LDL-C (mg/dl) | 142.2 ± 33.9 | 136.3 ± 32.2 | 139.7 ± 33.3 | 0.103 |
| TG (mg/dl) | 123.0 (100.5) | 89.4 (51.0) | 105.0 (76.9) | <0.001 |
| **Markers of glucose metabolism** |  |  |  |  |
| Fasting glucose (mg/dl) | 106.9 ± 23.5 | 98.1 ± 16.5 | 103.2 ± 21.3 | <0.001 |
| Fasting insulin (mU/ml) | 11.8 (7.8) | 9.3 (5.4) | 8.8 (7.4) | <0.001 |
| HbA1c (%) | 5.56 ± 0.83 | 5.51 ± 0.49 | 5.54 ± 0.71 | 0.540 |
| 2-hour insulin (μU/ml) <sup>b</sup> | 46.0 (70.5) | 42.0 (39.5) | 44.0 (51.8) | 0.412 |
| 2-hour glucose (mg/dl) <sup>b</sup> | 117.4 ± 44.5 | 104.0 ± 32.9 | 111.6 ± 40.4 | 0.003 |
| Diabetes status |  |  |  | 0.002 |
| Diabetes | 16.0 % (33) | 6.8 % (10) | 12.2 % (43) |  |
| Prediabetes | 26.7 % (55) | 19.0 % (28) | 23.5 % (83) |  |
| Normoglycemic | 57.3 % (118) | 74.1 % (109) | 64.3 % (227) |  |
| **Markers of renal function** |  |  |  |  |
| Glomerular filtration rate (ml/min/1.73m<sup>2</sup>) | 93.6 ± 16.7 | 91.2 ± 16.9 | 92.6 ± 16.8 | 0.195 |
| Uric acid (mg/dl) | 6.33 ± 1.32 | 4.57 ± 1.11 | 5.60 ± 1.51 | <0.001 |
| Creatinine (mg/dl) | 0.96 ± 0.13 | 0.77 ± 0.12 | 0.88 ± 0.16 | 0.001 |
| Albumin (g/dl) | 4.41 ± 0.29 | 4.28 ± 0.27 | 4.35 ± 0.29 | <0.001 |
| Cystatin C (mg/l) | 0.89 ± 0.14 | 0.85 ± 0.17 | 0.88 ± 0.16 | 0.029 |
| Urine albumin (mg/l) | 6.48 (10.52) | 6.22 (8.47) | 6.32 (8.85) | 0.431 |
| Urine creatinine (g/l) | 1.75 ± 0.74 | 1.39 ± 0.82 | 1.60 ± 0.79 | <0.001 |
| **Complete blood count** |  |  |  |  |
| Haematocrit (l/l) | 0.43 ± 0.03 | 0.39 ± 0.03 | 0.42 ± 0.03 | <0.001 |
| Thrombocytes (/nl) | 221.1 ± 51.6 | 244.5 ± 52.2 | 230.9 ± 53.1 | <0.001 |
| Erythrocytes (/pl) | 4.87 ± 0.37 | 4.45 ± 0.37 | 4.70 ± 0.40 | <0.001 |
| Leucocytes (/nl) | 5.61 (1.93) | 5.66 (1.98) | 5.65 (1.91) | 0.576 |
| Haemoglobin (g/l) | 150.4 ± 10.1 | 134.8 ± 9.8 | 143.9 ± 12.6 | <0.001 |

| Electrolyte panel |  |  |  |  |
| --- | --- | --- | --- | --- |
| Potassium (mmol/l) | 4.32 ± 0.31 | 4.22 ± 0.22 | 4.28 ± 0.28 | 0.001 |
| Sodium (mmol/l) | 139.0 (4.0) | 139.0 (3.5) | 139.0 (4.0) | 0.448 |
| Magnesium (mmol/l) | 0.85 ± 0.08 | 0.87 ± 0.08 | 0.86 ± 0.07 | 0.062 |
| Phosphate (mmol/l) | 0.98 ± 0.13 | 1.12 ± 0.14 | 1.04 ± 0.15 | <0.001 |
| Blood pressure parameters |  |  |  |  |
| SBP (mmHg) | 125.5 ± 16.0 | 113.0 ± 14.9 | 120.3 ± 16.4 | <0.001 |
| DBP (mmHg) | 77.6 ± 10.3 | 71.8 ± 8.2 | 75.3 ± 9.9 | <0.001 |
| Hypertension | 37.4 % (77) | 25.9 % (38) | 32.6 % (115) | 0.025 |
| Liver parameters |  |  |  |  |
| GGT (U/l) | 35.3 (33.9) | 19.6 (17.5) | 27.0 (28.0) | <0.001 |
| AST (U/l) | 24.5 (9.0) | 20.0 (8.0) | 23.0 (9.0) | <0.001 |
| ALT (U/l) | 31.0 (15.8) | 21.00 (12.0) | 27.0 (17.0) | <0.001 |
| Hepatic iron (s<sup>-1</sup>) | 41.8 ± 4.7 | 39.2 ± 4.1 | 40.7 ± 4.6 | <0.001 |
| Right liver lobe (s<sup>-1</sup>) | 42.4 ± 5.4 | 39.7 ± 4.3 | 41.3 ± 5.2 | <0.001 |
| Left liver lobe (s<sup>-1</sup>) | 41.1 ± 5.4 | 38.7 ± 4.7 | 40.1 ± 5.2 | <0.001 |
| Hepatic fat fraction (%) | 7.02 (10.08) | 3.53 (4.28) | 5.38 (7.92) | <0.001 |
| Right liver lobe (%) | 7.78 (9.99) | 3.96 (4.72) | 6.10 (8.99) | <0.001 |
| Left liver lobe (%) | 6.39 (10.62) | 3.16 (4.34) | 4.53 (7.63) | <0.001 |
| Further laboratory values |  |  |  |  |
| Alkaline phosphatase (U/l) | 65.9 ± 17.9 | 67.6 ± 23.6 | 66.6 ± 20.5 | 0.448 |
| CRP (mg/l) | 1.09 (1.70) | 1.26 (2.00) | 1.12 (1.78) | 0.349 |
| Vitamin D (ng/ml) | 24.3 ± 11.8 | 22.2 ± 11.3 | 23.4 ± 11.6 | 0.094 |
| Troponin T (pg/ml) | 3.55 (5.20) | 1.50 (0.77) | 1.50 (3.82) | <0.001 |
| Behavioral risk factors |  |  |  |  |
| Alcohol consumption (g/day) | 20.1 (36.5) | 3.1 (12.8) | 8.6 (25.9) | <0.001 |
| Smoking status |  |  |  | 0.243 |
| Smoker | 19.9% (41) | 22.4% (33) | 21.0% (74) |  |
| Ex-smoker | 46.1% (95) | 35.4% (52) | 41.6% (147) |  |
| Never-smoker | 34.0% (70) | 42.2% (62) | 37.4% (132) |  |
| Pack years<sup>c</sup> | 17.8 (28.7) | 10.9 (17.0) | 15.2 (23.2) | <0.001 |
| Physical activity |  |  |  | 0.046 |
| Active | 55.3% (114) | 66.0% (97) | 59.8% (211) |  |
| Inactive | 44. % (92) | 34.0% (50) | 40.2% (142) |  |
| Medication intake |  |  |  |  |
| Beta-blockers | 11.7% (24) | 11.6% (17) | 11.6% (41) | 1.000 |
| ACE inhibitors | 8.7% (18) | 13.6% (20) | 10.8% (38) | 0.169 |
| Calcium antagonists | 6.8% (14) | 7.5% (11) | 7.1% (25) | 0.822 |
| Diuretics | 11.7% (24) | 13.6% (20) | 12.5% (44) | 0.620 |
| Antihypertensives | 23.3% (48) | 25.2% (37) | 24.1% (85) | 0.709 |
| Lipid-lowering agents | 10.2% (21) | 10.2% (15) | 10.2% (36) | 1.000 |
| Treatment of hyperuricemia | 4.4% (9) | 0% (0) | 2.5% (9) | 0.012 |

Values are given as arithmetic means ± standard deviation or median (IQR). Categorical variables are given as percentages (counts).
<sup>a</sup>p-values are from t-test or Mann-Whitney-U Test, and X<sup>2</sup> Test, respectively.
Abbreviations: SBP, systolic blood pressure; DBP, diastolic blood pressure; HDL-C, high-density lipoprotein cholesterol; LDL-C, low-density lipoprotein cholesterol; TG, triglycerides; GGT, gamma-glutamyl transferase; AST, aspartate aminotransferase; ALT, alanine aminotransferase; CRP, c-reactive protein; HFF, hepatic fat fraction.
<sup>b</sup>Considering only participants with 2-hour insulin/-glucose data from OGTT (N=323, 184 men, 139 women).
<sup>c</sup>Considering only ex-smokers and current smokers with available data (N=213, 130 men, 83 women).

Mean laboratory values were within the non-pathological range (for reference ranges, see Supplementary Table 3). For example, liver enzymes were within normal ranges for both men and women (GGT: men 35.3 U/l, women: 19.6 U/l; AST: men 24.5 U/l, women: 20.0 U/l; ALT: men 31.0 U/l, women 21.0 U/l).

### Distribution of HIC and Correlation with Age, HFF and Genetic Risk Score

HIC was significantly higher in men than in women (41.8 s^-1^ ± 4.7 and 39.2 s^-1^ ± 4.1, p <0.001 respectively). The distribution was approximately normal. Applying the cut-off value of R2^*^ > 41 s^-1^ defined by Kühn et al. (14). 44.5% of the participants would be diagnosed with mild hepatic iron overload. However, no participant had moderate to severe iron overload (cutoffs 62.5 s^-1^ and 70.1 s^-1^, respectively). Age was significantly correlated with HIC in women (rho=0.48, p<0.001) but not in men (rho=0.11, p=0.13, see Figure 2). HFF was correlated with HIC in both men and women (rho=0.32, p<0.001, and rho=0.51, p<0.001, respectively, see Figure 2).

**Figure 2:**
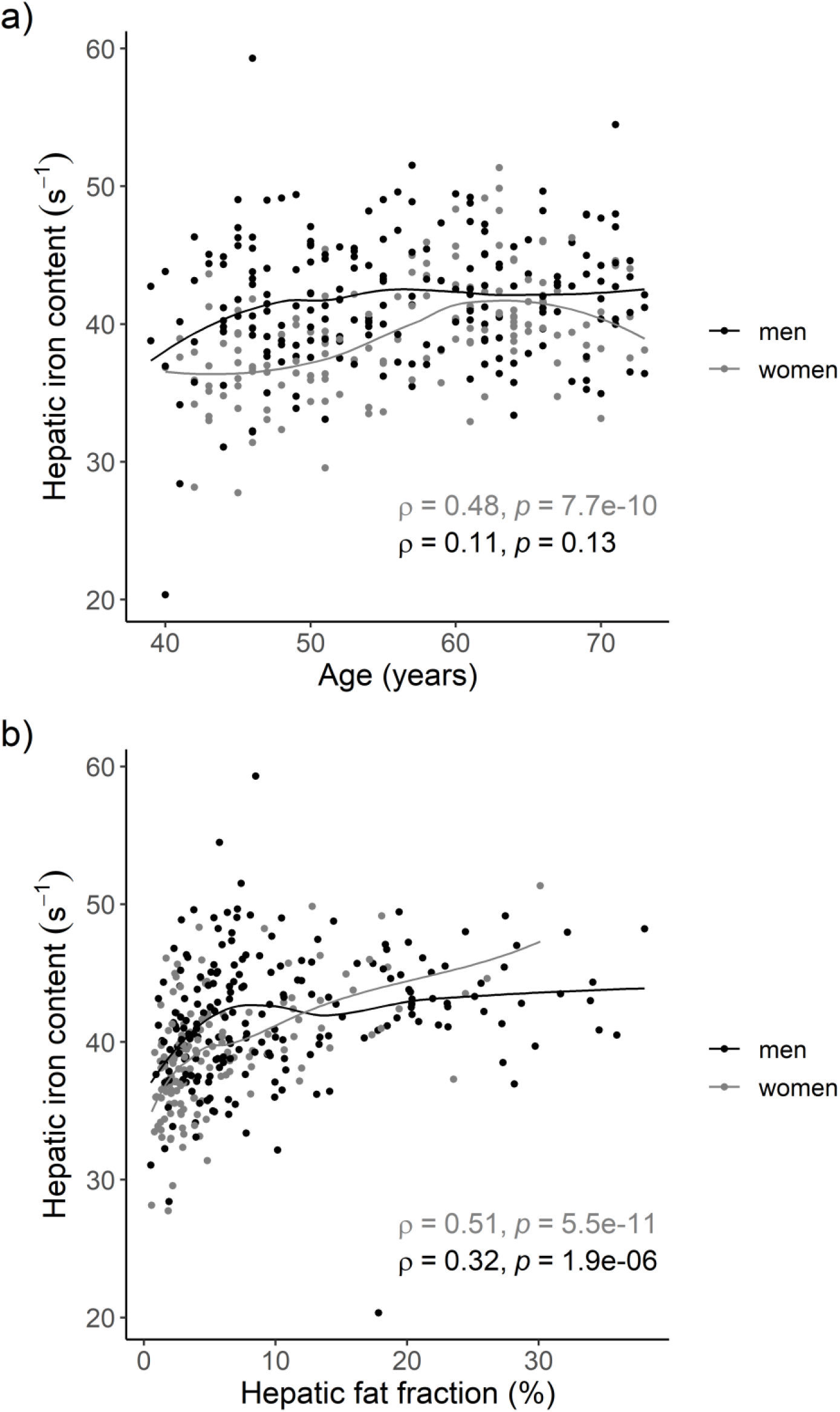
Scatter plots showing the sex-specific correlations of HIC with a) age and b) HFF, respectively. Lines denote the regression lines derived from locally weighted smoothing. Rho denotes the Spearman’s rank correlation coefficients.

Using a cutoff of HFF ≥ 5.6 %, 129 men (62.6%) and 44 women (29.9%) had hepatic steatosis. HIC was higher in individuals with hepatic steatosis compared to those without (men: 42.8 s^-1^ vs 40.0 s^-1^, p<0.001, women: 41.7 s^-1^ vs 38.1 s^-1^, p<0.001) (Supplementary Figure 1).

HIC increased with quartiles of the genetic risk score (Figure 3), resulting in significant differences in HIC between the lowest and highest quartile (men: 40.6 s^-1^ vs 43.1 s^-1^, p=0.007, women: 37.7 s^-1^ vs 40.2 s^-1^, p=0.03).

**Figure 3:**
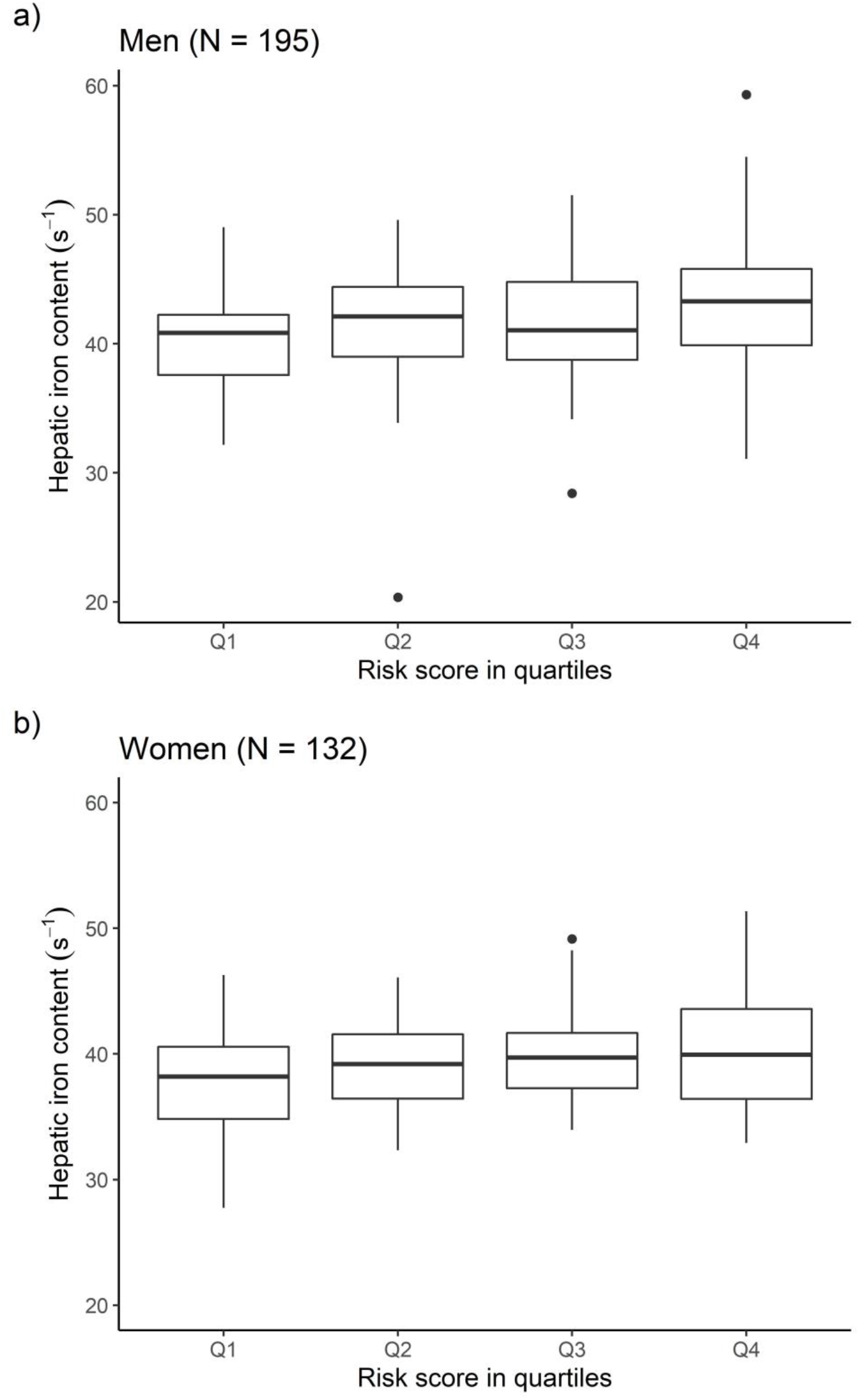
Boxplots of HIC according to genetic risk score quartiles for a) men and b) women.

### Identification of Relevant Associated Variables

Relevant factors associated with HIC identified by LASSO regression are depicted in Figure 4. For men, most frequently selected variables were HFF, HbA1c and prediabetes, whereas for women, most frequently selected variables were age, HFF and VAT. Alcohol consumption was selected in both men and women. When excluding HFF from the analysis, results were mainly stable. Further selected parameters included fasting insulin, uric acid, triglycerides, vitamin D and beta-blocker use (Supplementary Figure 2).

**Figure 4:**
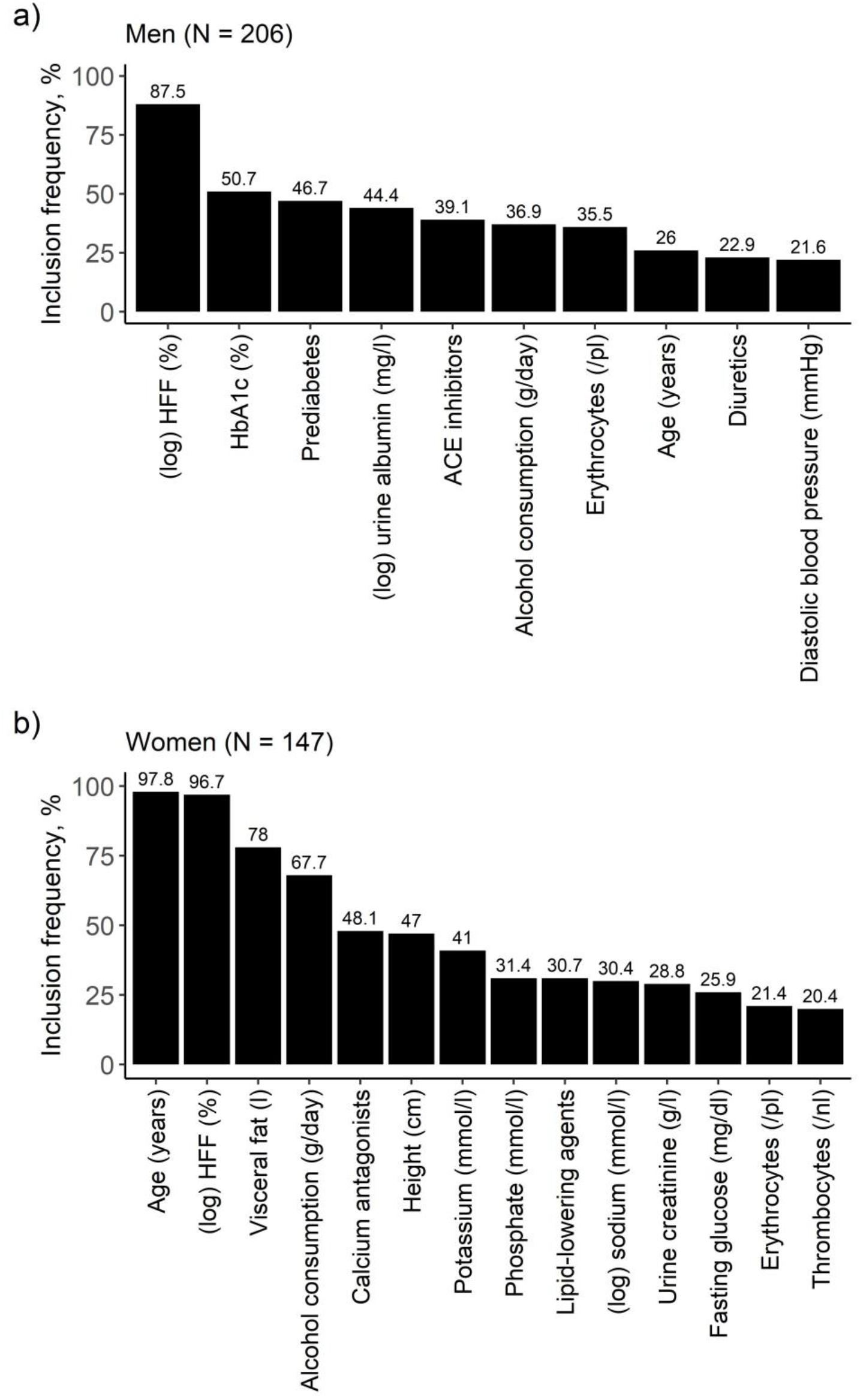
Bar diagrams of results from the model including HFF for a) men and b) women. Relevant variables were identified by variable selection through LASSO regression on 1000 bootstrap samples. On the y-axis: Inclusion frequency of the respective variable across 1000 bootstrap samples. Presented are only variables with an inclusion frequency > 20%.

In a sensitivity analysis including only participants who underwent an OGTT (N=323), results remained largely stable, but 2-hour glucose and 2-hour insulin were additionally selected as relevant covariates (Supplementary Figures 3, 4).

In the genetic analyses, selected variables also remained mostly the same (Figure 5). Leucocytes were further selected for both sexes and the genetic risk score was among the most frequently selected variables.

**Figure 5:**
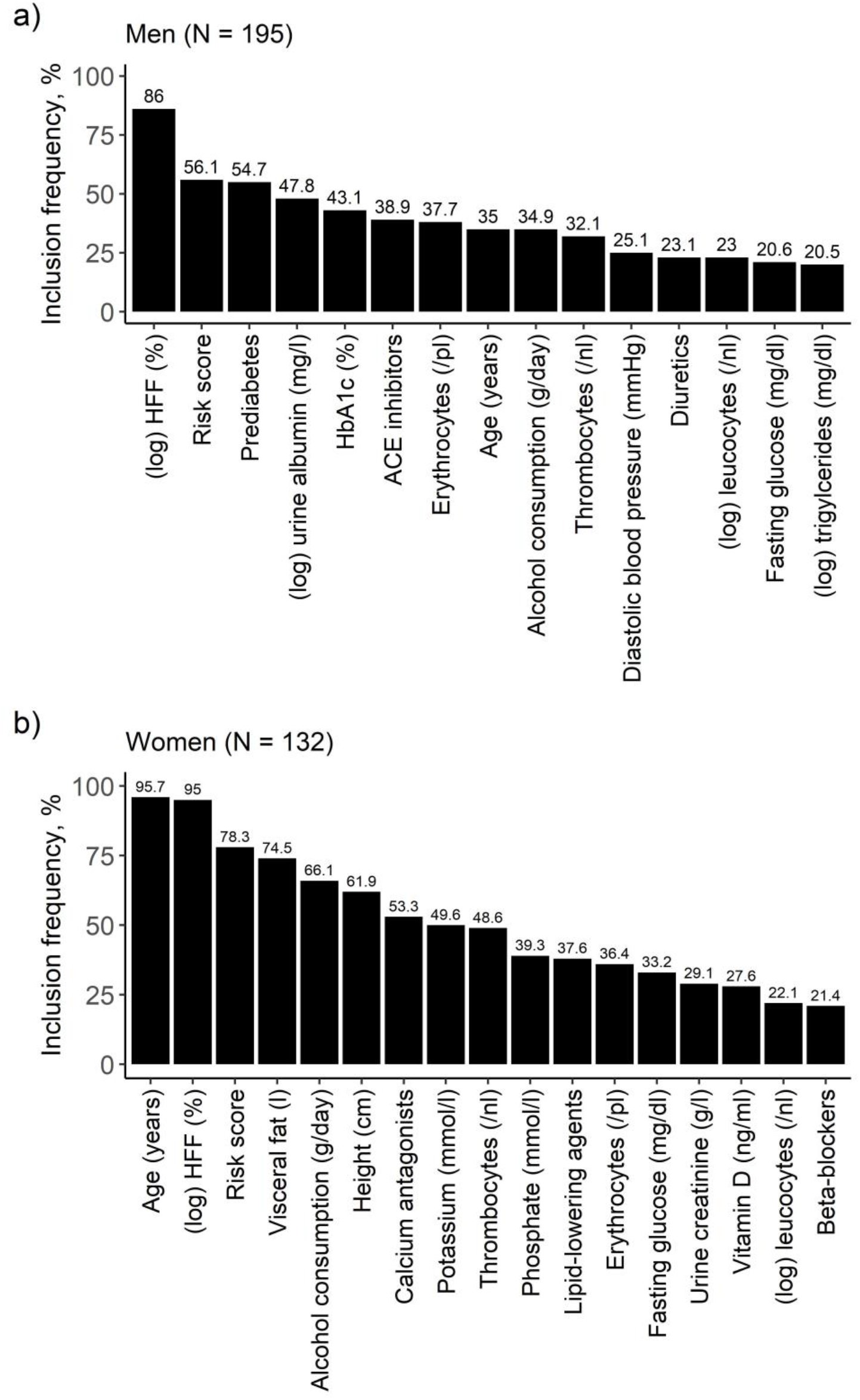
Bar diagrams of results from the model including genetic risk score for a) men and b) women. Relevant variables were identified by variable selection through LASSO regression on 1000 bootstrap samples. On the y-axis: Inclusion frequency of the respective variable across 1000 bootstrap samples. Presented are only variables with an inclusion frequency > 20%.

### Strength of Effects

Table 2 and 3 show the results of unpenalized linear regression analyses with and without adjustment for HFF for all variables that were identified in LASSO regression.

**Table 2:** Men: Results from unpenalized linear regression analyses. β denotes the regression coefficient of the respective variable for outcome HIC. Adjusted R^2^ denotes the variance of HIC explained. Presented are only variables with an inclusion frequency >20% in the variable selection procedure.

| | Adjustment | $\beta$ | 95% CI | p-value | adjusted $R^2$ |
| --- | --- | --- | --- | --- | --- |
| **Blood lipids** |  |  |  |  |  |
| (log) triglycerides (mg/dl) | age + HFF | 0.25 | -0.94 ; 1.44 | 0.68 | 0.08 |
|  | age | 1.11 | -0.01 ; 2.22 | 0.05 | 0.03 |
| **Markers of glucose metabolism** |  |  |  |  |  |
| (log) fasting insulin (mU/ml) | age + HFF | -0.42 | -1.73 ; 0.9 | 0.53 | 0.08 |
|  | age | 0.97 | -0.14 ; 2.07 | 0.09 | 0.02 |
| HbA1c (%) | age + HFF | -1.44 | -2.17 ; -0.71 | 0.00 | 0.14 |
|  | age | -1.11 | -1.87 ; -0.36 | 0.00 | 0.05 |
| Prediabetes | age + HFF | 0.92 | -0.67 ; 2.5 | 0.25 | 0.11 |
|  | age | 2.13 | 0.64 ; 3.63 | 0.01 | 0.05 |
| **Markers of renal function** |  |  |  |  |  |
| Uric acid (mg/dl) | age + HFF | 0.23 | -0.27 ; 0.72 | 0.37 | 0.08 |
|  | age | 0.49 | 0.01 ; 0.97 | 0.05 | 0.03 |
| (log) urine albumin (mg/l) | age + HFF | -0.80 | -1.29 ; -0.31 | 0.00 | 0.12 |
|  | age | -0.69 | -1.2 ; -0.19 | 0.01 | 0.04 |
| **Complete blood count** |  |  |  |  |  |
| Thrombocytes (/nl) | age + HFF | -0.01 | -0.02 ; 0.01 | 0.38 | 0.08 |
|  | age | -0.01 | -0.02 ; 0 | 0.20 | 0.02 |
| Erythrocytes (/pl) | age + HFF | -1.35 | -3.05 ; 0.35 | 0.12 | 0.09 |
|  | age | -1.03 | -2.79 ; 0.73 | 0.25 | 0.02 |
| **Blood pressure** |  |  |  |  |  |
| Systolic blood pressure (mmHg) | age + HFF | 0.01 | -0.03 ; 0.06 | 0.48 | 0.08 |
|  | age | 0.04 | 0 ; 0.08 | 0.05 | 0.03 |
| Diastolic blood pressure (mmHg) | age + HFF | 0.04 | -0.02 ; 0.11 | 0.17 | 0.09 |
|  | age | 0.08 | 0.02 ; 0.14 | 0.01 | 0.04 |
| **Liver parameters** |  |  |  |  |  |
| (log) hepatic fat fraction (%) | age | 1.46 | 0.74 ; 2.19 | 0.00 | 0.08 |
| **Behavioral risk factors** |  |  |  |  |  |
| Alcohol consumption (g/day) | age + HFF | 0.02 | 0 ; 0.05 | 0.04 | 0.10 |
|  | age | 0.03 | 0.01 ; 0.05 | 0.01 | 0.04 |
| **Medication intake** |  |  |  |  |  |
| ACE inhibitors | age + HFF | -3.61 | -5.79 ; -1.43 | 0.00 | 0.12 |
|  | age | -2.78 | -5.03 ; -0.53 | 0.02 | 0.04 |
| Diuretics | age + HFF | -2.50 | -4.44 ; -0.56 | 0.01 | 0.11 |
|  | age | -1.97 | -3.97 ; 0.04 | 0.05 | 0.03 |
| **Genetic analyses (N=195)** |  |  |  |  |  |
| Genetic risk score, continuous | age + HFF | 0.64 | 0.16 ; 1.12 | 0.01 | 0.12 |

CI, confidence interval; HFF, hepatic fat fraction.

**Table 3:** Women: Results from unpenalized linear regression analyses. β denotes the regression coefficient of the respective variable for outcome HIC. Adjusted R^2^ denotes the variance of HIC explained. Presented are only variables with an inclusion frequency >20% in the variable selection procedure.

| | Adjustment | $\beta$ | 95% CI | p-value | adjusted $R^2$ |
| --- | --- | --- | --- | --- | --- |
| **Body composition** |  |  |  |  |  |
| Height (cm) | age + HFF | -0.04 | -0.13 ; 0.05 | 0.40 | 0.36 |
|  | age | -0.05 | -0.15 ; 0.05 | 0.29 | 0.21 |
| Visceral fat (l) | age + HFF | 0.01 | -0.12 ; 0.14 | 0.91 | 0.35 |
|  | age | 0.81 | 0.5 ; 1.13 | 0.00 | 0.33 |
| **Blood lipid markers** |  |  |  |  |  |
| (log) triglycerides (mg/dl) | age + HFF | -0.20 | -1.8 ; 1.4 | 0.81 | 0.35 |
|  | age | 1.70 | 0.14 ; 3.26 | 0.03 | 0.23 |
| **Markers of glucose metabolism** |  |  |  |  |  |
| Fasting glucose (mg/dl) | age + HFF | 0.01 | -0.02 ; 0.05 | 0.47 | 0.36 |
|  | age | 0.05 | 0.01 ; 0.09 | 0.01 | 0.24 |
| **Markers of renal function** |  |  |  |  |  |
| Urine creatinine (g/l) | age + HFF | -0.47 | -1.14 ; 0.21 | 0.17 | 0.36 |
|  | age | -0.14 | -0.89 ; 0.6 | 0.70 | 0.21 |
| **Complete blood count** |  |  |  |  |  |
| Thrombocytes (/nl) | age + HFF | 0.01 | -0.01 ; 0.02 | 0.34 | 0.36 |
|  | age | 0.00 | -0.01 ; 0.01 | 0.71 | 0.21 |
| Erythrocytes (/pl) | age + HFF | -1.24 | -3.02 ; 0.55 | 0.17 | 0.36 |
|  | age | -0.91 | -2.9 ; 1.07 | 0.36 | 0.21 |
| **Electrolyte panel** |  |  |  |  |  |
| Potassium (mmol/l) | age + HFF | -2.74 | -5.13 ; -0.34 | 0.03 | 0.38 |
|  | age | -2.44 | -5.11 ; 0.22 | 0.07 | 0.22 |
| (log) sodium (mmol/l) | age + HFF | -19.63 | -45.67 ; 6.4 | 0.14 | 0.36 |
|  | age | -40.47 | -67.31 ; -13.63 | 0.00 | 0.25 |
| Phosphate (mmol/l) | age + HFF | 2.62 | -1.51 ; 6.76 | 0.21 | 0.36 |
|  | age | 0.61 | -3.94 ; 5.15 | 0.79 | 0.21 |
| **Liver parameters** |  |  |  |  |  |
| (log) hepatic fat fraction (%) | age | 2.08 | 1.38 ; 2.79 | 0.00 | 0.36 |
| **Further laboratory values** |  |  |  |  |  |
| Vitamin D (ng/ml) | age + HFF | -0.02 | -0.07 ; 0.03 | 0.46 | 0.36 |
|  | age | -0.04 | -0.1 ; 0.01 | 0.10 | 0.22 |
| **Behavioral risk factors** |  |  |  |  |  |
| Alcohol consumption (g/day) | age + HFF | 0.05 | 0.02 ; 0.09 | 0.00 | 0.39 |
|  | age | 0.07 | 0.03 ; 0.11 | 0.00 | 0.26 |
| **Medication intake** |  |  |  |  |  |
| Beta-blockers | age + HFF | 0.31 | -1.53 ; 2.15 | 0.74 | 0.35 |
|  | age | 1.69 | -0.26 ; 3.63 | 0.09 | 0.22 |
| Calcium antagonists | age + HFF | -2.24 | -4.29 ; -0.19 | 0.03 | 0.37 |
|  | age | -2.35 | -4.63 ; -0.07 | 0.04 | 0.23 |
| Lipid-lowering agents | age + HFF | 1.19 | -0.67 ; 3.06 | 0.21 | 0.36 |
|  | age | 1.89 | -0.14 ; 3.93 | 0.07 | 0.22 |
| **Genetic analyses (N=132)** |  |  |  |  |  |
| Genetic risk score,<br>continuous | age + HFF | 0.65 | 0.16; 1.14 | 0.01 | 0.39 |

CI, confidence interval; HFF, hepatic fat fraction.

In general, associations attenuated after adjustment for HFF. Variance of outcome explained (adjusted R^2^) was generally higher in women (21-39%) than in men (2-14%).

In men, of all variables identified by LASSO regression HFF, HbA1c, urine albumin, alcohol consumption, ACE inhibitors, and diuretics were also significantly associated with HIC in unpenalized regression. Higher values of HbA1c were negatively associated with HIC (β=-1.44, p<0.001), whereas higher consumption of alcohol was associated with increased HIC (β=0.02, p=0.04). Urine albumin and diuretics showed a negative relationship with HIC (β=-0.80, p<0.01 and β=-2.50, p=0.01, respectively).

In women, age, HFF, potassium, alcohol consumption, and calcium antagonists were also significantly associated with HIC in unpenalized regression. We revealed a negative relationship between HIC and potassium and calcium antagonist intake, respectively (β=-2.74, p=0.03 and β=-2.24, p=0.03). Alcohol consumption was also associated with increased HIC (β=0.05, p<0.01).

The continuous genetic risk score was positively associated with HIC in both men and women (β=0.64, p<0.01 and β=0.65, p<0.01, respectively).

## Discussion

In this explorative study, we investigated sex-specific distributions of HIC in a population-based sample and identified associated factors from a large panel of markers. Overall, HIC was normally distributed with significantly lower values in women and none of the participants exceeds the threshold for severe hepatic iron overload. We revealed notable sex-specific associations of HIC with markers of body composition, glucose metabolism and alcohol consumption.

### Distribution of HIC and Effect of Age

The distribution of HIC in our sample was comparable to other studies. Kühn et al. (14) reported median HIC values of 34.4 s^-1^ and the UK Biobank (15) found mean values of 44.02 s^-1^ compared to our 40.7 s^-1^. Kühn et al.’s cutoff suggested that 44.5% of the participants of the current study present mild iron overload. As expected, this is a higher prevalence than found by Kühn et al. (17.4%) and a slightly lower prevalence than in the UK Biobank study (51.5%). We found higher values of HIC in men compared to women, which is coherent with the aforementioned studies (14, 15). Higher levels of HIC in men might be explained by higher levels of testosterone since androgens are known to be regulators of hepcidin expression (24). Besides, men had higher HFF levels than women, and HFF is substantially associated with HIC, as outlined below. Moreover, most women before onset of menopause regularly excrete iron through menstrual bleeding, leading to generally lower body iron. We found a strong correlation between age and HIC in women, suggesting a link to the onset of menopause. Our findings regarding the effect of age on HIC are supported by the study of Obrzut et al. (25), who examined considerably younger participants and reported distinctly lower HIC levels (mean: 28.7 s^-1^).

### Body Composition and Blood Lipid Markers

We identified HFF as a main associated factor with HIC. This is in line with results from the UK Biobank (15) and MRI studies on patient samples (26, 7). In our study, the association was stronger in women than in men.

Furthermore, our results demonstrated that VAT is associated with HIC in women. This may be explained by the fact that the iron-regulating hormone hepcidin is expressed in abdominal adipose tissue, in addition to the liver which is the main site of synthesis (27). Consequently, higher amounts of adipose tissue stimulate the HAMP gene and increase hepcidin production. This pathway is independent of diabetes status (27), which might explain the stability of VAT as an associated factor with HIC among all analyses.

Our procedure selected triglycerides as a relevant factor for both sexes. This finding is supported by the observation of Jehn et al. (28), who reported a significant increase in serum ferritin with increasing triglyceride levels. In addition, a study including only participants with iron overload due to hemochromatosis reported elevated triglyceride levels as well (29). Related to this finding is the selection of lipid-lowering agents in women, which might serve as a proxy for underlying hypertriglyceridemia in this context.

In summary, our results indicate a relationship between abdominal adipose tissue and lipid profile with hepatic iron storage. This relationship is more pronounced in women.

### Genetic Effects

The genetic risk score was frequently selected as a relevant associated factor with HIC. Weights of the respective SNPs were notably different between men and women (Supplementary Table 2), indicating sex-specific effects. Genetic variants rs1799945 and rs1800562 in HFE showed the strongest association with HIC in men and women, respectively. Both SNPs lead to hepatic iron overload due to decreased hepcidin levels (30, 31) and are additionally associated with ferritin and transferrin (32, 33). Moreover, variants rs855791 and rs4820268 in TMPRSS6 are known to be associated with iron traits including transferrin, serum iron and ferritin (34, 30), since TMPRSS6 modulates the transcription of hepcidin (34). A mendelian randomization study analysing UK Biobank data revealed a causal relationship between central obesity and elevated HIC (30). It is hypothesized that an interplay between genetics and dietary factors and a cross-talk between liver and adipose tissue is responsible for the causal effect of abdominal obesity on HIC.

### Markers of Glucose Metabolism

Several diabetes-related markers were selected to be associated with HIC, even after exclusion of participants with established T2DM. We found an association with HIC for prediabetes and HbA1c in men, which is in line with Britton et al. (9) who found an inverse correlation between HIC and HbA1c. On the other hand, Kühn et al. (14) reported that HbA1c was not a relevant predictor of iron overload in their study. Recent findings from a subcohort of the aforementioned study showed a stronger association between serum ferritin and T2DM and an altered glucose metabolism even in the absence of pathologic iron overload, suggesting a combined effect of hepatic iron overload and ferritin (35). The relationship between prediabetes and increased HIC is consistent with other studies, analysing the association of diabetes status and serum ferritin levels (36, 12).

Our results regarding fasting glucose and HIC are conflicting since we found a positive association between HIC and fasting glucose in women but a negative association in men, whereas 2-hour glucose showed a positive association only in men. Animal studies showed an increase in blood glucose levels in animals with iron overload, indicating that increased iron storage might be associated with altered glucose metabolism (37). A mendelian randomization study analysing UK Biobank data revealed a potentially causal association of fasting glucose with increased HIC (30).

An association between iron and diabetes risk in hereditary iron metabolism disorders such as hemochromatosis is already established (38). Even nonpathologically increased body iron stores are related with higher risks for development of T2DM (37). Haap et al. (39) found a positive association between HIC with T2DM and insulin resistance. Moreover, dysmetabolic iron overload syndrome (DIOS), defined as the presence of iron overload and insulin resistance, is frequently observed in patients with MetS (37). Consequently, there seems to be an association between insulin-resistance syndrome and iron overload (12). Increased ROS are observed in iron deficiency as well as iron overload syndromes and ROS are known to induce beta cell damage and insulin resistance (1). An overactivation of gluconeogenesis leading to increased hepcidin expression is discussed as a pathway, leading to iron accumulation and cell damage within the liver. This indicates an interplay between hepatic dysfunction, serum ferritin and metabolic disorders. Our results therefore confirm and expand previous findings regarding the association of markers of glucose metabolism with HIC.

### Alcohol Consumption

The link between alcohol consumption and HFF is already established (40). Our results also show that a higher consumption of alcohol is associated with increased HIC, independent of HFF. These results are consistent with Whitfield et al. (41) who reported that even moderate alcohol consumption raises body iron stores. Furthermore, patients with alcoholic liver disease have been found to show alcohol induced suppression of hepcidin. Alcohol induces hypoxia, which is known to reduce the expression of hepatic HAMP leading to decreased hepcidin levels (42).

### Renal Function Parameters and Diuretics

The data-driven approach revealed uric acid as a relevant associated factor with HIC in men and women. Previous studies including healthy adults, reported a positive correlation between serum ferritin and uric acid independent of gender and age (43). Furthermore, another study reported a worsening of hepatic and renal functioning when a simultaneous elevation in uric acid and serum ferritin levels were present (44). Potential mechanisms of the association between iron overload with increased uric acid could be related to oxidative stress or insulin sensitivity. Additionally, our group previously found an association between increased uric acid and HFF (45).

Contrary to formerly reported positive relations between serum ferritin and proteinuria (46), we found a negative association between HIC and urine albumin in men when including participants with established diabetes in the analysis. The results might differ due to the heterogeneous study populations, since Kim et al. (46) excluded patients with diabetes from the analyses.

We found that diuretic use was associated with lower HIC with relatively large effect sizes. Diuretics are frequently prescribed in patients with renal diseases and one study described a high proportion of anaemia in haemodialysis patients, which in turn was associated with increased inflammatory status (47). Systemic inflammation leads to an upregulation of signal transducers and activators of transcription 3 (STAT3) and increases the synthesis of hepcidin followed by decrease in iron levels (48). This pathway is as a possible explanation for our findings, indicating a role of renal function markers in liver iron storage.

### Complete Blood Count

Selection of erythrocytes as a relevant variable was stable among the different models for both sexes, relating increased levels of erythrocytes with decreased HIC. We speculate that an increase in erythrocyte levels mirrors the expansion of erythropoiesis due to an increased iron demand within the body. To sufficiently cover the demand, hepcidin expression is suppressed and iron stored within the liver is released (49). Interestingly, haemoglobin, the iron containing protein in erythrocytes, and haematocrit, were not among the selected variables associated with HIC in our study, whereas Kühn et al. (14) revealed mean corpuscular haemoglobin as the most predictive marker for HIC. Additionally, we demonstrated that the selection of thrombocytes was more frequent among women compared to men. Thrombocytopenia is associated with iron deficiency due to the increased risk for haemorrhages and another study reports a correlation between HIC and thrombocytes in patients with transfusion-related iron overload (50).

### Electrolyte Panel and Medication

We identified potassium as a relevant marker in women associated with a decrease in HIC. Given that iron overload is associated with T2DM, this relationship is plausible since hypokalaemia is associated with an increased risk for T2DM due to reduced insulin sensitivity (51). However, diuretic use can also affect potassium balance and, as mentioned above, diuretic use was also found to be associated with HIC.

Additionally, we observed a negative association between sodium and HIC in women. Hyponatremia is frequently observed in cirrhotic patients and decreased serum levels correlate with severity of cirrhosis (52). Our results indicate that this association might already be visible in the non-pathological range.

Use of cardiovascular medication (ACE inhibitors in men, calcium antagonists in women) was found to be relevantly associated with decreased HIC. Associations of cardiovascular medication with serum ferritin have already been suggested (53) but conclusive findings about the effect of antihypertensive medication on iron metabolism are lacking. Results from animal studies suggest that a decrease in divalent metal transporter-1 (DMT-1) expression due to calcium antagonists may be responsible for a reduction in iron absorption (54).

### Strength and Limitations

Out study has unique strengths. The study sample from an established population-based cohort was well characterized which enabled the analysis of a rich set of markers and risk factors. The assessment of HIC by MRI allowed for a precise quantification of both hepatic iron and fat content. Moreover, we applied appropriate statistical techniques to identify relevant associated factors and ensured robustness and stability of our findings.

Nevertheless, our study has several limitations which need to be addressed. Most importantly, we lacked data of serum indices of iron metabolism, such as ferritin and hepcidin. Further research on disentangling the association of circulating iron markers and markers of iron storage is necessary. Moreover, the available data set was limited to a relatively small size. Therefore, replication and extension of our findings in larger population-based cohorts are needed. One opportunity is the German National Cohort, a population-based study within Germany with MRI data on 30000 participants, which would enable more intricate analyses with higher statistical power.

## Conclusion

Our results indicate sex-specific associations of MRI-derived HIC with several factors, specifically markers of glucose metabolism, renal function, body composition, alcohol intake and genetic markers. Thus, our study extends previous knowledge of relevant HIC-related factors to a population-based sample. Further work is required to disentangle the complexity of pathways between disorders of iron homeostasis and pathologies.

## Supporting information

Supplementary_Distributionandassociatedfactorsofhepaticiron

## Data Availability

Restrictions apply to the availability of some or all data generated or analysed during this study to preserve patient confidentiality or because they were used under license. The corresponding author will on request detail the restrictions and any conditions under which access to some data may be provided.

## Acknowledgements

The authors are grateful for the participants who volunteered to provide the data. We thank the field workers, radiologists, interviewers, technicians, and computer assistants for their contributions to the collection of data.

## Author Contributions

LM performed the statistical analyses, evaluated the results and drafted the manuscript. FB and AP conceived and designed the KORA-MRI study. RvK, JN, FB and CLS collected the MRI data and analyzed the images. RvK, RL, JN, FB and CLS contributed substantially to MRI data preparation and quality assurance. BT, WK, WR and AP contributed substantially to the collection, quality assurance and data preparation of the biomarker measurements within KORA. RvK, RL, JF, JN, BT, WK, WR, FB, CLS, and AP contributed substantially to the scientific content and interpretation of the results. SR conceived the study question, contributed to the statistical analyses, the interpretation of the results and the drafting of the manuscript. SR had primary responsibility for final content. All authors read and approved the final manuscript.

