## Supplementary_Distributionandassociatedfactorsofhepaticiron for "Distribution and associated factors of hepatic iron – a population-based imaging study"

**Supplementary Table 1:** Description of variable assessment.

| Variable | Description |
|---|---|
| **Body composition** | |
| Body weight (kg) | Measured by calibrated steelyards or digital scales (SECA 635 or SECA 877 or SECA measuring station 285, Seca GmbH & Co, KG, Hamburg, Germany) |
| Height (cm) | Measured by by calibrated levelling bar (SECA 242, Seca GmbH & Co, KG, Hamburg, Germany) |
| BMI (kg/m$^2$) | Calculated by dividing weight in kilogram by squared height in meter |
| Waist circumference (cm) | Measured with an inelastic tape at the level midway between the lower rib margin and the iliac crest |
| Hip circumference (cm) | Measured with an inelastic tape at the level of maximal gluteal protrusion |
| Subcutaneous fat (l) | MRI measurement (3 Tesla): subcutaneous fat from the femoral head to the cardiac apex by a three-dimensional in/opposed-phase VIBE-Dixon sequence |
| Visceral fat (l) | MRI measurement (3 Tesla): visceral fat from the femoral head to the diaphragm by a three-dimensional in/opposed-phase VIBE-Dixon sequence |
| Total fat (l) | MRI measurement (3 Tesla): total fat from the femoral head to the cardiac apex by a three-dimensional in/opposed-phase VIBE-Dixon sequence |
| **Blood lipids** | |
| Total cholesterol (mg/dl) | Enzymatic, colorimetric CHOL Flex assay (Vista, Siemens or Cobas, Roche) |
| HDL-C (mg/dl) | Enzymatic, colorimetric HDLC Flex assay (Vista, Siemens or Cobas, Roche) |
| LDL-C (mg/dl) | Enzymatic, colorimetric LDLC Flex assay (Vista, Siemens or Cobas, Roche) |
| TG (mg/dl) | Enzymatic, colorimetric TRIG Flex assay (Vista, Siemens or Cobas, Roche) |
| **Markers of glucose metabolism** | |
| Fasting glucose (mg/dl) | UV test using enzymatic reference method with hexokinase (Vista, Siemens or Cobas, Roche) |
| Fasting insulin (mU/ml) | Elecsys Insulin immunoassay with two monoclonal antibodies (Vista, Siemens or Cobas, Roche) |
| HbA1c (%) | Cation-exchange high performance liquid chromatographic, photometric assay (VARIANT II TURBO Hemoglobin Testing System, Bio-Rad Laboratories Inc, Hercules, US) |
| 2-hour insulin (µU/ml) | Serum insulin 2 hours post OGTT |
| 2-hour glucose (mg/dl) | Serum glucose 2 hours post OGTT |
| Diabetes status | Established type 2 diabetes mellitus or results from OGTT |
| Categories: | |
| *Normoglycemic* | 2-hour glucose <140 mg/dL and fasting glucose level <110 mg/dL |
| *Prediabetes* | 2-hour glucose between 140 and 200 mg/dL and/or an fasting glucose between 110 and 125 mg/dL, and normal 2-hour glucose |
| *Diabetes* | 2-hour glucose >200 mg/dL and/or fasting glucose >125 mg/dL (1) |

| | |
|---|---|
| **Markers of renal function** | |
| Glomerular filtration rate (calculated from cystatin C) | Sex-specific calculation based on serum creatinine according to CKD-EPI |
| Glomerular filtration rate (calculated from cystatin C and serum creatinine) | Sex-specific calculation based on serum creatinine according to CKD-EPI |
| Uric acid (mg/dl) | Enzymatic colorimetric UA Flex assay (Vista, Siemens or Cobas, Roche) |
| Creatinine (mg(dl) | Kinetic colorimetric CREJ assay based on Jaffé method |
| Albumin (g/dl) | Bromocresol purple (BCP) dye-binding method with ALB Flex assay |
| Urine albumin (mg/l) | Immunonephelometry |
| Urine creatinine (g/l) | Kinetic colorimetric CREJ assay based on Jaffé method |
| Cystatin C (mg/l) | Particle-enhanced immunonephelometry |
| **Complete blood count** | |
| Thrombocytes (/nl) | Impedance measures according to Beckman-Coulter method |
| Erythrocytes (/pl) | Impedance measures according to Beckman-Coulter method |
| Leucocytes (/nl) | Impedance measures according to Beckman-Coulter method |
| Haemoglobin (g/l) | Impedance measures according to Beckman-Coulter method |
| Haematocrit (l/l) | Impedance measures according to Beckman-Coulter method |
| **Electrolyte panel** | |
| Potassium (mmol/l) | Flame photometry, spectrophotometry, direct or indirect ion selective electrode potentiometry |
| Sodium (mmol/l) | Flame photometry, spectrophotometry, direct or indirect ion selective electrode potentiometry |
| Magnesium (mmol/l) | Modified methylthymol blue complexometric procedure including Ba-EGTA |
| Phosphate (mmol/l) | Modified phosphomolybdate method including p-methylaminophenol sulfate and bisulfite |
| **Blood pressure** | |
| Systolic blood pressure (mmHg) | 3 measurements with an oscillometric digital device (OMRON HEM-705CP). Average of 2nd and 3rd measurements |
| Diastolic blood pressure (mmHg) | 3 measurements with an oscillometric digital device (OMRON HEM-705CP). Average of 2nd and 3rd measurements |
| Hypertension | Blood pressure ≥ 140/90 or use of antihypertensive medication given that participants were aware of having hypertension |
| **Liver parameters** | |
| GGT (U/l) | Modified IFCC method:including L-gamma-glutamyl-3-carboxy-4-nitranilide with glycylglycine |
| AST (U/l) | Modified IFCC method: P5Ü as activator and lactic acid dehydrogenase (LDH) to eliminate pyruvate interference |
| ALT (U/l) | Modified IFCC method: P5Ü as activator and hydroxymethyl aminomethane as buffer |
| Hepatic iron, right and left liver lobe ($s^{-1}$) | MRI measurement (3 Tesla) by a single-voxel spectroscopy with a high-speed T2-corrected multi-echo (HISTO) technique |

| | |
|---|---|
| HFF, right and left liver lobe (%) | MRI measurement (3 Tesla) by a multiecho single-voxel 1H spectroscopy |
| **Further laboratory values** | |
| Alkaline phosphatase (U/l) | Bowers+McComb method: change in absorbance at 405 nm due to the formation of p-nitrophenol (p-NP) in the presence of the transphosphorylating buffer and AMP |
| CRP (mg/L) | CardioPhase hsCRP by particle enhanced immunonephelometry |
| Vitamin D (ng/ml) | Enhanced chemiluminescence immunoassay |
| Troponin T (pg/ml) | ECLIA Immunologic test |
| **Behavioral risk factors** | |
| Age (years) | Self-reported in standardized interview |
| Alcohol consumption (g/day) | Self-reported in standardized interview, calculated based on reported amount and type of beverages consumed, further information Bayerl et al. (2) |
| Smoking status | Self-reported in standardized interview, further information Bayerl et al.(2) |
| Pack years | calculated based on reported number of cigarettes smoked, further information Bayerl et al. (2) |
| Physically active | Self-reported in standardized interview, further information Rabel et al. (3) |
| **Medication intake** | |
| All medications[a] | Assessed by standardized interview. Participants were asked to bring packages of their medications from the last 7 days before the interview, further information Teuner et al. (4) |

[a]please not that although the reference might not pertain to KORA FF4 but to one of the other KORA surveys, the described procedure was also applicable in FF4.

**Supplementary Table 2:** SNPs included in the genetic risk score.

| Marker of iron metabolism | SNP | Position | Gene | Alt Allele | MAF KORA | Weight, men | Weight, women | Ref |
|---|---|---|---|---|---|---|---|---|
| unsaturated iron binding capacity, total iron binding capacity | rs2698530 | 2:64503895 | MIR4433B | C | 0.25 | -0.165 | -0.194 | McLaren (5) |
| Ferritin | rs744653 | 2:190378750 | SLC40A1 | T | 0.16 | -0.736 | 0.596 | Meidtner (6) |
| Ferritin | rs5742933 | 2:190649316 | PMS1 | C | 0.20 | 0.495 | 0.480 | Liao (7) |
| Transferrin | rs1799852 | 3:133475722 | TF | T | 0.11 | -1.152 | 0.016 | He (8) |
| Transferrin, total iron binding capacity | rs3811647 | 3:133484029 | TF | A | 0.33 | 0.099 | 0.040 | McLaren (5) He (8) Tayrac (9) |
| Ferritin, Hepcidin, Liver Iron | rs1799945 | 6:26091179 | HFE | G | 0.15 | 1.204 | -0.273 | Meidtner (6) He (8) Wilman (10) Yuan (11) |
| Ferritin, Hepcidin, total iron binding capacity, Liver Iron | rs1800562 | 6:26093141 | HFE | A | 0.05 | 1.876 | 3.269 | McLaren (5) Meidtner (6) He (8) Wilman (10) Yuan (11) Traglia (12) |
| Transferrin | rs4841132 | 8:9183596 | PPP1R3B | G | 0.08 | 0.162 | 1.133 | Raffield (13) |
| Ferritin, soluble transferrin receptor (sTfR) | rs236918 | 11:117091609 | PCSK7 | C | 0.11 | 0.947 | 0.220 | Meidtner (6) Oexle (14) |
| Ferritin, Hepcidin, soluble transferrin receptor (sTfR), Liver Iron | rs855791 | 22:37462936 | TMPRSS6 | G | 0.43 | -0.619 | 0.296 | Meidtner (6) He (8) Wilman (10) Yuan (11) Traglia (12) Oexle (14) Nai (15) Wainaina (16) |
| Hepcidin | rs4820268 | 22:37469591 | TMPRSS6 | A | 0.45 | -0.251 | 0.192 | Wainaina (16) |
| Serum iron | rs1421312 | 22:37487810 | TMPRSS6 | G | 0.38 | -0.167 | 0.434 | McLaren (17) |
| Ferritin | rs738409 | 22:44324727 | PNPLA3 | G | 0.24 | -0.027 | 0.374 | Hotta (18) |

MAF: minor allele frequency. Sex-specific weights were calculated from univariate regression of the respective SNP on outcome HIC.

**Supplementary Table 3:** Reference intervals of laboratory values, with corresponding mean, minimum and maximum values in the study sample.

| | Unit | Reference range | Ref | Study sample mean ± SD or median (IQR) | Study sample min | Study sample max |
|---|---|---|---|---|---|---|
| **Blood lipids** | | | | | | |
| Total cholesterol | mg/dl | < 240 | (19) | 218.1 ± 36.7 | 133.0 | 347.0 |
| HDL-C | mg/dl | > 40 | (20) | 62.1 ± 17.7 | 25.0 | 112.0 |
| LDL-C | mg/dl | < 160 | (20) | 139.7 ± 33.3 | 50.0 | 230.0 |
| Triglycerides | mg/dl | < 150 | (20) | 105.0 (76.9) | 32.0 | 628.3 |
| **Markers of glucose metabolism** | | | | | | |
| Fasting glucose | mg/dl | 70–110 | (19) | 103.2 ± 21.3 | 77.0 | 305.0 |
| Fasting insulin | mU/ml | < 25 | (19) | 8.8 ± 7.4 | 0.1 | 54.0 |
| HbA1c | % | < 5.7 | (21) | 5.54 ± 0.71 | 0.04 | 13.6 |
| **Markers of renal function** | | | | | | |
| Glomerular filtration rate | ml/min/1.73m$^2$ | < 90 | (22) | 92.6 ± 16.8 | 29.5 | 122.8 |
| Uric acid | mg/dl | m: 3.5 - 7.0<br>w: 2.5 - 6.5 | (19) | m: 6.33 ± 1.32<br>w: 4.57 ± 1.11 | 2.15 | 11.76 |
| Creatinine | mg/dl | m: 0.84 - 1.25<br>w: 0.66 - 1.09 | (19) | m: 0.96 ± 0.13<br>w: 0.77 ± 0.12 | 0.55 | 1.26 |
| Albumine | g/dl | 3.5 - 5.3 | (19) | 4.35 ± 0.29 | 3.57 | 5.03 |
| Cystatin C | mg/l | m: 0.54 - 0.94<br>w: 0.45 - 0.82 | (19) | m: 0.89 ± 0.14<br>w: 0.85 ± 0.17 | 0.55 | 1.96 |
| Urine albumine | mg/l | < 20 | (23) | 6.32 (8.85) | 1.10 | 1070.0 |
| Urine creatinine | g/l | 0.3 - 2.5 | (24) | 1.60 ± 0.79 | 0.10 | 4.12 |
| **Complete blood count** | | | | | | |
| Haematocrit | l/l | m: 0.40 - 0.52<br>w: 0.35 - 0.47 | (19) | m: 0.43 ± 0.03<br>w: 0.39 ± 0.03 | 0.29 | 0.50 |
| Thrombocytes | /nl | 150 - 450 | (19) | 230.9 ± 53.1 | 89.0 | 455.0 |
| Erythrocytes | /pl | m: 4.40 - 5.90<br>w. 3.80 - 5.20 | (19) | m: 4.87 ± 0.37<br>w: 4.45 ± 0.37 | 3.56 | 5.80 |
| Leucocytes | /nl | 4.30 - 10.0 | (19) | 5.65 (1.91) | 2.50 | 15.02 |
| Haemoglobin | g/l | m: 133 - 177<br>w: 117 - 157 | (19) | m: 150.4 ± 10.1<br>w: 134.8 ± 9.8 | 94.0 | 179.0 |
| **Electrolyte panel** | | | | | | |
| Potassium | mmol/l | 3.5 - 5.1 | (19) | 4.28 ± 0.28 | 2.92 | 6.13 |
| Sodium | mmol/l | 135 -145 | (19) | 139.0 (4.0) | 121.0 | 145.0 |
| Magnesium | mmol/l | 0.65 - 1.05 | (19) | 0.86 ± 0.07 | 0.24 | 1.01 |
| Phosphate | mmol/l | 0.87 - 1.67 | (19) | 1.04 ± 0.15 | 0.53 | 1.49 |
| **Liver parameters** | | | | | | |
| GGT | U/l | m: < 60<br>w: < 40 | (19) | m: 35.3 (33.9)<br>w: 19.6 (17.5) | 7.0 | 348.0 |
| AST | U/l | m: < 50<br>w: < 35 | (20) | m: 24.5 (9.0)<br>w: 20.0 (8.0) | 7.0 | 184.0 |
| ALT | U/l | m: < 50<br>w: < 35 | (20) | m: 31.0 (15.8)<br>w: 21.0 (12.0) | 9.0 | 123.0 |

| Further laboratory values | | | | | | |
|---|---|---|---|---|---|---|
| Alkaline phosphatase | U/l | m: 40 - 130<br>w: 35 - 105 | (19) | m: 65.9 ± 17.9<br>w: 67.6 ± 23.6 | 20.0 | 212.0 |
| CRP | mg/l | < 5 | (19) | 1.12 (1.78) | 0.16 | 25.0 |
| Vitamin D | ng/ml | < 20 | (25) | 23.4 ± 11.6 | 4.2 | 64.0 |
| hs-Troponin T | pg/ml | < 14 | (26) | 1.50 (3.82) | 1.50 | 29.1 |

**Supplementary Table 4:** Men: Results from unpenalized linear regression analyses from sensitivity analysis. β denotes the regression coefficient of the respective variable for outcome HIC. Adjusted $R^2$ denotes the variance of HIC explained. Presented are only variables with an inclusion frequency >20% in the variable selection procedure.

| | Adjustment | β | 95% CI | p-value | adjusted $R^2$ |
|---|---|---|---|---|---|
| **Body composition** | | | | | |
| Waist circumference (cm) | age + HFF | -0.07 | -0.14 ; 0 | 0.04 | 0.10 |
| | age | 0.02 | -0.04 ; 0.07 | 0.49 | 0.01 |
| **Blood lipids** | | | | | |
| (log) trigylcerides (mg/dl) | age + HFF | 0.25 | -0.94 ; 1.44 | 0.68 | 0.08 |
| | age | 1.11 | -0.01 ; 2.22 | 0.05 | 0.03 |
| **Markers of glucose metabolism** | | | | | |
| Fasting glucose (mg/dl) | age + HFF | -0.05 | -0.07 ; -0.02 | 0.00 | 0.13 |
| | age | -0.03 | -0.06 ; 0 | 0.04 | 0.03 |
| (log) 2-hour insulin (µU/ml) | age + HFF | 0.06 | -0.73 ; 0.86 | 0.88 | 0.12 |
| | age | 1.01 | 0.33 ; 1.69 | 0.00 | 0.05 |
| 2-hour glucose (mg/dl) | age + HFF | 0.01 | -0.01 ; 0.03 | 0.20 | 0.13 |
| | age | 0.02 | 0.01 ; 0.04 | 0.00 | 0.06 |
| Prediabetes | age + HFF | 0.92 | -0.67 ; 2.5 | 0.25 | 0.11 |
| | age | 2.13 | 0.64 ; 3.63 | 0.01 | 0.05 |
| **Complete blood count** | | | | | |
| Thrombocytes (/nl) | age + HFF | -0.01 | -0.02 ; 0.01 | 0.38 | 0.08 |
| | age | -0.01 | -0.02 ; 0 | 0.20 | 0.02 |
| Erythrocytes (/pl) | age + HFF | -1.35 | -3.05 ; 0.35 | 0.12 | 0.09 |
| | age | -1.03 | -2.79 ; 0.73 | 0.25 | 0.02 |
| **Electrolyte panel** | | | | | |
| (log) sodium (mmol/l) | age + HFF | 4.63 | -19.6 ; 28.86 | 0.71 | 0.08 |
| | age | -2.89 | -27.69 ; 21.92 | 0.82 | 0.01 |
| Magnesium (mmol/l) | age + HFF | -2.40 | -10.23 ; 5.43 | 0.55 | 0.08 |
| | age | -2.22 | -10.33 ; 5.9 | 0.59 | 0.01 |
| **Blood pressure** | | | | | |
| Diastolic blood pressure (mmHg) | age + HFF | 0.04 | -0.02 ; 0.11 | 0.17 | 0.09 |
| | age | 0.08 | 0.02 ; 0.14 | 0.01 | 0.04 |
| **Liver parameters** | | | | | |
| (log) hepatic fat fraction (%) | age | 1.46 | 0.74 ; 2.19 | 0.00 | 0.08 |
| **Behavioral risk factors** | | | | | |
| Alcohol consumption (g/day) | age + HFF | 0.02 | 0 ; 0.05 | 0.04 | 0.10 |
| | age | 0.03 | 0.01 ; 0.05 | 0.01 | 0.04 |
| **Medication intake** | | | | | |
| Calcium antagonists | age + HFF | -2.27 | -4.77 ; 0.23 | 0.07 | 0.09 |
| | age | -2.28 | -4.87 ; 0.31 | 0.08 | 0.02 |

CI: confidence interval, HFF: hepatic fat fraction.

**Supplementary Table 5:** Women: Results from unpenalized linear regression analyses from sensitivity analysis. β denotes the regression coefficient of the respective variable for outcome HIC. Adjusted $R^2$ denotes the variance of HIC explained. Presented are only variables with an inclusion frequency >20% in the variable selection procedure.

| | Adjustment | β | 95% CI | p-value | adjusted $R^2$ |
|---|---|---|---|---|---|
| **Body composition** | | | | | |
| Height (cm) | age + HFF | -0.04 | -0.13 ; 0.05 | 0.40 | 0.36 |
| | age | -0.05 | -0.15 ; 0.05 | 0.29 | 0.21 |
| Visceral fat (l) | age + HFF | 0.01 | -0.12 ; 0.14 | 0.91 | 0.35 |
| | age | 0.81 | 0.5 ; 1.13 | 0.00 | 0.33 |
| **Blood lipids** | | | | | |
| Total cholesterol (mg/dl) | age + HFF | 0.01 | -0.01 ; 0.02 | 0.45 | 0.36 |
| | age | 0.01 | -0.01 ; 0.03 | 0.46 | 0.21 |
| (log) trigylcerides (mg/dl) | age + HFF | -0.20 | -1.8 ; 1.4 | 0.81 | 0.35 |
| | age | 1.70 | 0.14 ; 3.26 | 0.03 | 0.23 |
| **Markers of glucose metabolism** | | | | | |
| Fasting glucose (mg/dl) | age + HFF | 0.01 | -0.02 ; 0.05 | 0.47 | 0.36 |
| | age | 0.05 | 0.01 ; 0.09 | 0.01 | 0.24 |
| (log) fasting insulin (mU/ml) | age + HFF | -0.09 | -1.13 ; 0.94 | 0.86 | 0.35 |
| | age | 1.35 | 0.43 ; 2.27 | 0.00 | 0.25 |
| (log) 2-hour insulin (µU/ml) | age + HFF | -1.15 | -2.21 ; -0.1 | 0.03 | 0.35 |
| | age | 0.56 | -0.41 ; 1.54 | 0.25 | 0.20 |
| **Markers of renal function** | | | | | |
| Uric acid (mg/dl) | age + HFF | -0.01 | -0.6 ; 0.57 | 0.96 | 0.35 |
| | age | 0.73 | 0.18 ; 1.28 | 0.01 | 0.24 |
| Urine creatinine (g/l) | age + HFF | -0.47 | -1.14 ; 0.21 | 0.17 | 0.36 |
| | age | -0.14 | -0.89 ; 0.6 | 0.70 | 0.21 |
| **Complete blood count** | | | | | |
| Thrombocytes (/nl) | age + HFF | 0.01 | -0.01 ; 0.02 | 0.34 | 0.36 |
| | age | 0.00 | -0.01 ; 0.01 | 0.71 | 0.21 |
| Erythrocytes (/pl) | age + HFF | -1.24 | -3.02 ; 0.55 | 0.17 | 0.36 |
| | age | -0.91 | -2.9 ; 1.07 | 0.36 | 0.21 |
| **Electrolyte panel** | | | | | |
| Potassium (mmol/l) | age + HFF | -2.74 | -5.13 ; -0.34 | 0.03 | 0.38 |
| | age | -2.44 | -5.11 ; 0.22 | 0.07 | 0.22 |
| (log) sodium (mmol/l) | age + HFF | -19.63 | -45.67 ; 6.4 | 0.14 | 0.36 |
| | age | -40.47 | -67.31 ; -13.63 | 0.00 | 0.25 |
| Phosphate (mmol/l) | age + HFF | 2.62 | -1.51 ; 6.76 | 0.21 | 0.36 |
| | age | 0.61 | -3.94 ; 5.15 | 0.79 | 0.21 |
| **Liver parameters** | | | | | |
| (log) hepatic fat fraction (%) | age | 2.08 | 1.38 ; 2.79 | 0.00 | 0.36 |
| **Further laboratory values** | | | | | |
| (log) CRP (mg/l) | age + HFF | -0.37 | -0.92 ; 0.18 | 0.19 | 0.36 |
| | age | 0.17 | -0.41 ; 0.75 | 0.57 | 0.21 |
| Vitamin D (ng/ml) | age + HFF | -0.02 | -0.07 ; 0.03 | 0.46 | 0.36 |

| | | | | | |
|---|---|---|---|---|---|
| | age | -0.04 | -0.1 ; 0.01 | 0.10 | 0.22 |
| **Behavioral risk factors** | | | | | |
| Alcohol consumption (g/day) | age + HFF | 0.05 | 0.02 ; 0.09 | 0.00 | 0.39 |
| | age | 0.07 | 0.03 ; 0.11 | 0.00 | 0.26 |
| **Medication intake** | | | | | |
| Calcium antagonists | age + HFF | -2.24 | -4.29 ; -0.19 | 0.03 | 0.37 |
| | age | -2.35 | -4.63 ; -0.07 | 0.04 | 0.23 |
| Lipid-lowering agents | age + HFF | 1.19 | -0.67 ; 3.06 | 0.21 | 0.36 |
| | age | 1.89 | -0.14 ; 3.93 | 0.07 | 0.22 |

CI: confidence interval, HFF: hepatic fat fraction.

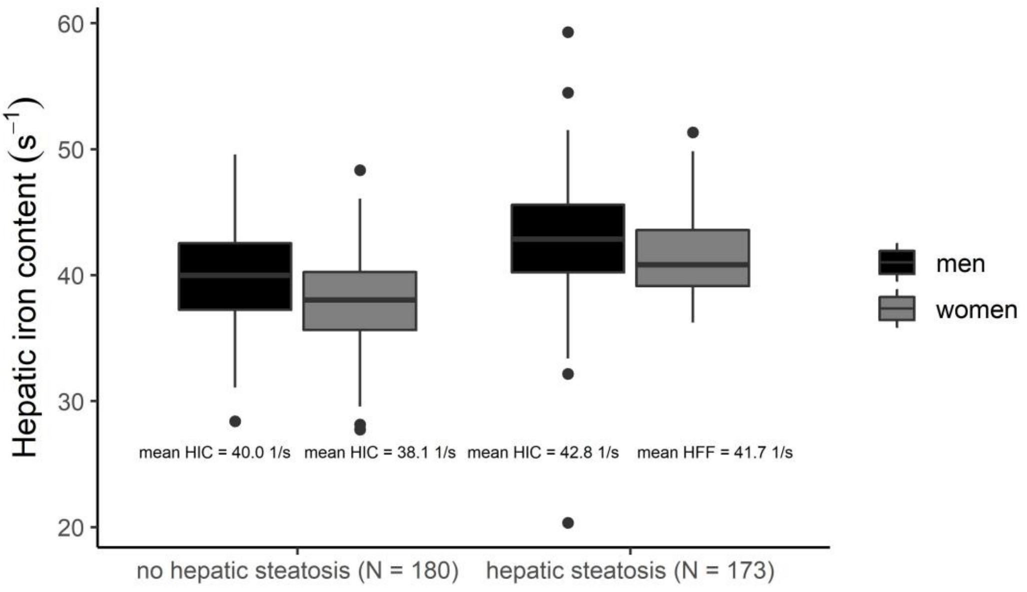


**Supplementary Figure 1:** Boxplots of HIC according to hepatic steatosis status by sex. Cutoff for hepatic steatosis was HFF ≥ 5.6%, as defined by Schaapman et al. (27).

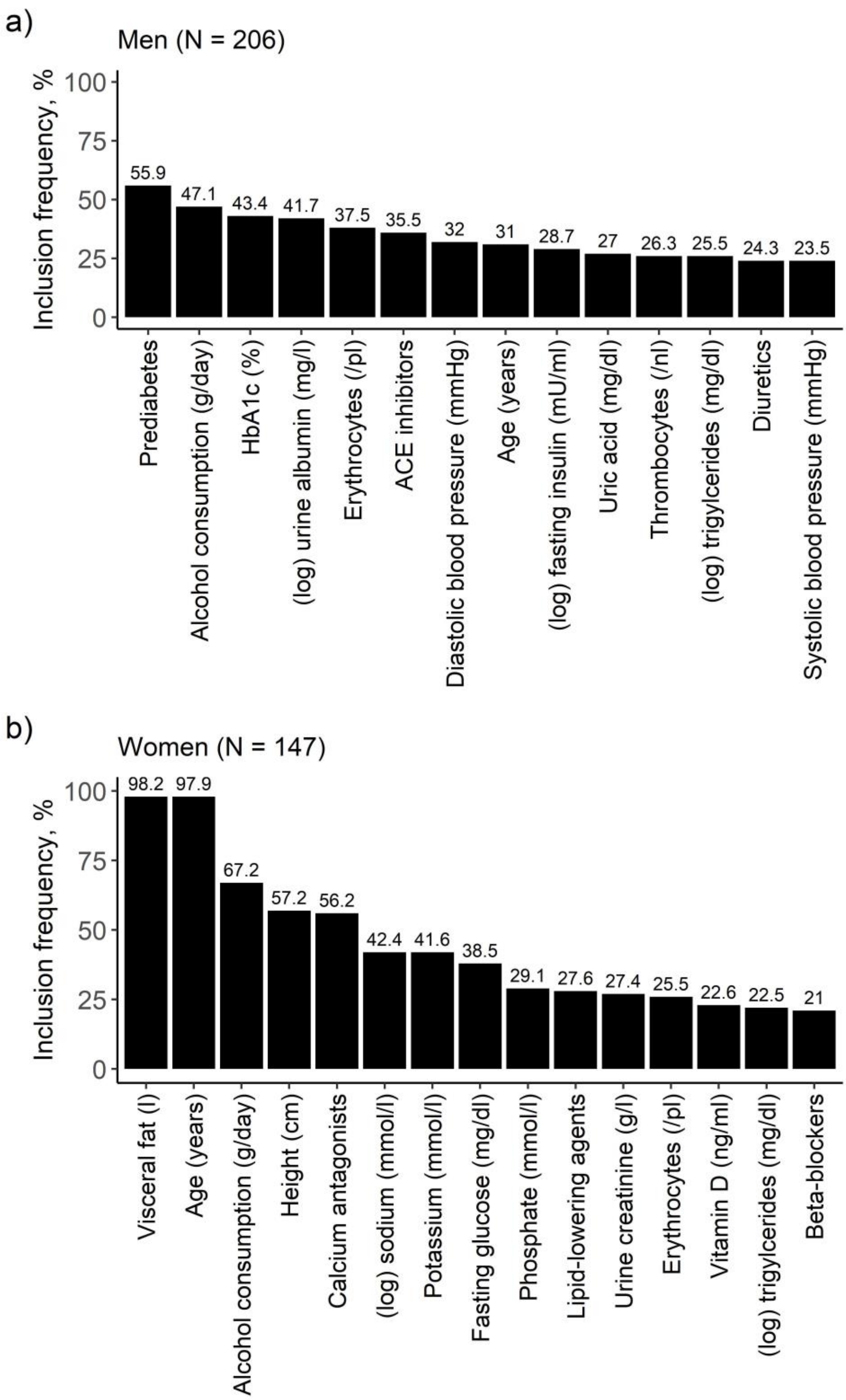


**Supplementary Figure 2:** Bar diagrams of results from the model excluding HFF for a) men and b) women. Relevant variables were identified by variable selection through LASSO regression on 1000 bootstrap samples. On the y-axis: Inclusion frequency of the respective variable across 1000 bootstrap samples. Presented are only variables with an inclusion frequency > 20%.

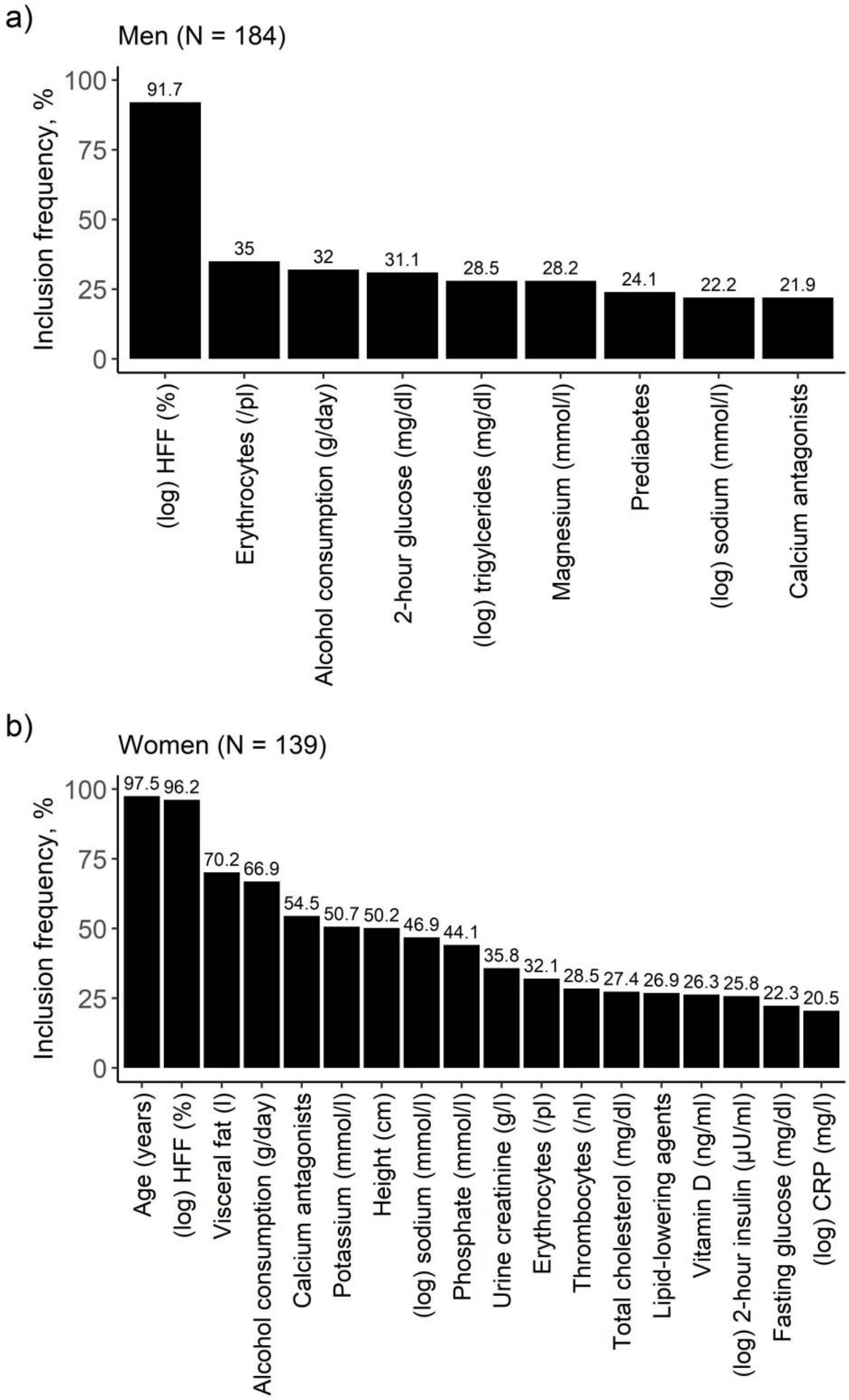


**Supplementary Figure 3:** Bar diagrams of results from the sensitivity analysis including HFF for a) men and b) women. Relevant variables were identified by variable selection through LASSO regression on 1000 bootstrap samples. On the y-axis: Inclusion frequency of the respective variable across 1000 bootstrap samples. Presented are only variables with an inclusion frequency > 20%.

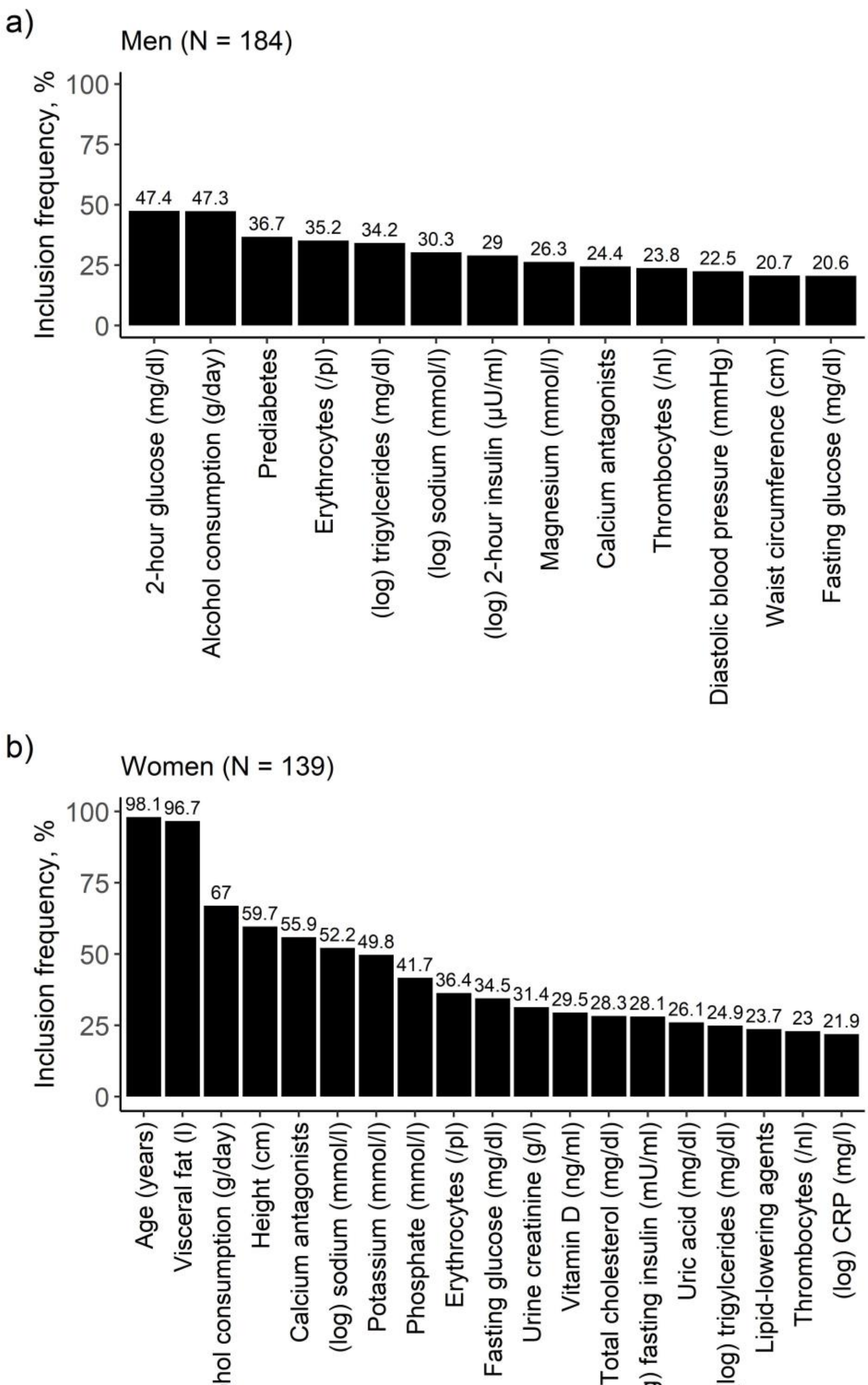


**Supplementary Figure 4:** Bar diagrams of results from the sensitivity analysis excluding HFF for a) men and b) women. Relevant variables were identified by variable selection through LASSO regression on 1000 bootstrap samples. On the y-axis: Inclusion frequency of the respective variable across 1000 bootstrap samples. Presented are only variables with an inclusion frequency > 20%.
